# Ethnic disparities in incident SARS-CoV-2 infections became wider during the second wave of SARS-CoV-2 in Amsterdam, the Netherlands: a population-based longitudinal study

**DOI:** 10.1101/2021.07.21.21260956

**Authors:** Liza Coyer, Anders Boyd, Janke Schinkel, Charles Agyemang, Henrike Galenkamp, Anitra D M Koopman, Tjalling Leenstra, Yvonne T H P van Duijnhoven, Eric P Moll van Charante, Bert-Jan H van den Born, Anja Lok, Arnoud Verhoeff, Aeilko H Zwinderman, Suzanne Jurriaans, Karien Stronks, Maria Prins

## Abstract

**Background:** Surveillance data in high-income countries have reported more frequent SARS-CoV-2 diagnoses in ethnic minority groups. We examined the cumulative incidence of SARS-CoV-2 and its determinants in six ethnic groups in Amsterdam, the Netherlands.

**Methods:** We analyzed participants enrolled in the population-based HELIUS cohort, who were tested for SARS-CoV-2-specific antibodies and answered COVID-19-related questions between June 24-October 9, 2020 (after the first wave) and November 23, 2020-March 31, 2021 (during the second wave). We modeled SARS-CoV-2 incidence from January 1, 2020-March 31, 2021 using Markov models adjusted for age and sex. We compared incidence between ethnic groups over time and identified determinants of incident infection within ethnic groups.

**Findings:** 2,497 participants were tested after the first wave; 2,083 (83·4%) were tested during the second wave. Median age at first visit was 54 years (interquartile range=44-61); 56·6% were female. Compared to Dutch-origin participants (15·9%), cumulative SARS-CoV-2 incidence was higher in participants of South-Asian Surinamese (25·0%; adjusted hazard ratio [aHR]=1·66;95%CI=1·16-2·40), African Surinamese (28·9%;aHR=1·97;95%CI=1·37-2·83), Turkish (37·0%;aHR=2·67;95%CI=1·89-3·78), Moroccan (41·9%;aHR=3·13;95%CI=2·22-4·42), and Ghanaian (64·6%;aHR=6·00;95%CI=4·33-8·30) origin. Compared to those of Dutch origin, differences in incidence became wider during the second versus first wave for all ethnic minority groups (all p for interaction<0.05), except Ghanaians. Having household members with suspected SARS-CoV-2 infection, larger household size, and low health literacy were common determinants of SARS-CoV-2 incidence across groups.

**Interpretation:** SARS-CoV-2 incidence was higher in the largest ethnic minority groups of Amsterdam, particularly during the second wave. Prevention measures, including vaccination, should be encouraged in these groups.

**Funding:** ZonMw, Public Health Service of Amsterdam, Dutch Heart Foundation, European Union, European Fund for the Integration of non-EU immigrants.

## Introduction

Higher rates of SARS-CoV-2 diagnoses were observed in ethnic minority groups, in particular people of African and Asian descent, during the first wave of the SARS-CoV-2 epidemic in the United Kingdom, United States, and much of Europe.^1-4^ SARS-CoV-2 seroprevalence estimates in England and the United States continued to increase in individuals of African and Asian descent during late 2020 and early 2021.^5,6^ These disparities have been related to ethnic differences in household composition, occupations requiring use of public transportation, and increased exposure to crowded conditions.^1,2,7,8^ Structural barriers to testing and healthcare access, and socioeconomic deprivation among ethnic minorities have also been implicated as reasons for ethnic differences in SARS-CoV-2 diagnoses.^1,2,9,10^

Much of the insight into ethnic differences in SARS-CoV-2 diagnoses and its determinants have relied on more specific cross-sectional studies or routine surveillance data, the latter of which are subject to changing SARS-CoV-2 testing capacity and policy and varying proportions of asymptomatic infections. Although these studies have been helpful, they make it difficult to infer upon the individual risk of incident SARS-CoV-2 infection and the factors leading up to incident infection. Furthermore, information on specific ethnic groups is not frequently collected or too broad for many surveillance systems, leading to strong information bias. Dynamic changes in behaviour over time may also contribute to faster or slower SARS-CoV-2 incidence rates, which have not been studied within a closed study population to date.

Despite these limitations, this insight has been important for larger cities in Europe, such as the Dutch capital Amsterdam, where half of the population comprises inhabitants with a migration history, including people with foreign-born parents.^11^ Although previous research in other settings would suggest that SARS-CoV-2 seroprevalence should be higher in ethnic minority groups in Amsterdam, evidence from the first wave of the SARS-CoV-2 epidemic has indicated otherwise. A large population-based study conducted after the first wave of the SARS-CoV-2 epidemic found that only individuals of Ghanaian origin were at higher risk of past exposure to SARS-CoV-2, whereas individuals of South-Asian Surinamese, African Surinamese, Turkish, or Moroccan origin had a similar risk compared with individuals of Dutch origin.^12^

Given the potential for ethnic differences during the subsequent waves of the SARS-CoV-2 epidemic and the limitations from previous studies, we aimed to investigate whether SARS-CoV-2 incidence differed between individuals of South-Asian Surinamese, African Surinamese, Ghanaian, Turkish, Moroccan and Dutch origin living in Amsterdam, specifically during the first and second waves of the Dutch SARS-CoV-2 epidemic. Using longitudinal seroprevalence data nested within the large, population-based cohort study HEalthy Life in an Urban Setting (HELIUS),^13^ we compared the SARS-CoV-2 incidence between ethnic groups and identified determinants of incident infection within ethnic groups in Amsterdam, the Netherlands.

## Methods

### Study design and population

The HELIUS study is a multi-ethnic cohort study conducted in Amsterdam, the Netherlands, which focuses on the causes of potential ethnic disparities in cardiovascular disease, mental health, and infectious diseases. Detailed procedures have been previously described.^13^ Briefly, HELIUS includes persons of Dutch, South-Asian Surinamese, African Surinamese, Ghanaian, Moroccan, and Turkish origin, aged between 18 and 70 years at inclusion. A random sample of persons, stratified by ethnic origin, was taken from the municipality register of Amsterdam and subjects were invited to participate. Between January 2011 and December 2015, a total of 24,789 individuals were included.^13^ Participants filled in a questionnaire and underwent a physical examination during which biological samples were obtained. Ethical approval for the HELIUS study was obtained from the Academic Medical Center Ethical Review Board. All participants provided written informed consent.

Ethnicity was defined according to the country of birth of the participant and their parents.^13^ Participants were considered to be of non-Dutch ethnic origin if (i) they were born abroad and had at least one parent born abroad (first generation) or (ii) they were born in the Netherlands but both their parents were born abroad (second generation). Participants of Dutch origin were born in the Netherlands with both parents who were born in the Netherlands. Surinamese participants were further classified as African Surinamese, South-Asian Surinamese, and Javanese/other/unknown Surinamese, based on self-reporting.

HELIUS participants were randomly selected within each ethnic group and asked to participate in a seroprevalence substudy consisting of two visits. The first visit took place between June 24 and October 9, 2020 ^12^ and the second visit between November 23, 2020 and June 4, 2021. During both visits, serum samples were collected by venipuncture and stored at -20°C for SARS-CoV-2 antibody testing. Trained interviewers asked participants questions on uptake of COVID-19-related prevention measures (including SARS-CoV-2 vaccination status after the start of the Dutch COVID-19 national vaccination program on January 6, 2021), potential exposure, infection, symptoms, and disease.

### Outcomes

SARS-CoV-2-specific antibodies were determined using the WANTAI SARS-CoV-2 Ab enzyme-linked immunosorbent assay (ELISA) (Wantai Biological Pharmacy Enterprise Co., Beijing, China), according to the manufacturer’s instructions. This ELISA detects IgA, IgM, and IgG against the receptor binding domain of the S-protein of SARS-CoV-2.^14^

### Covariates

We used the following covariates: *from the baseline visit of the HELIUS study*– demographics (i.e. age, sex, ethnicity, migration generation), socio-economic factors (i.e. educational level, working status, occupational level, number of people living in household), access-to-healthcare indicators (i.e. proficiency with Dutch language and health literacy); *from the first visit of the seroprevalence study*– job setting, type of people living in household; *from each visit of the seroprevalence substudy*– suspecting household member/steady partner was infected, COVID-19 behaviors in the past week (i.e. number of times leaving the house, type of locations visited, number of visitors, frequency of using public transportation), travelled abroad (in 2020 or since first visit).

### Statistical analysis

We estimated SARS-CoV-2 incidence as a function of calendar time. Follow-up began on January 1, 2020 assuming that all participants were negative for SARS-CoV-2. Follow-up continued until the date of first substudy visit (for those lost to follow-up or vaccinated between visits) or second visit (if occurring on or prior to March 31, 2021). We chose to administratively censor follow-up on March 31, 2021 given the few participants with testing after this date. Equivocal test results were excluded from the analysis.

Since we only had observed measurements at specific time points and the true date of incident SARS-CoV-2 infection was unknown, we decided to model the transition of negative to positive SARS-CoV-2 antibody test over time. To this end, we used a time-homogenous, continuous-time, two-state Markov model, which allows estimation of incidence when the exact transition times are unknown.^15^ The positive state was an absorbing state, i.e. once participants were SARS-CoV-2 antibody positive, they remained in this state. We modelled transition intensities, interpreted as the instantaneous rates of a transition occurring (i.e. incidence), which depend on the probability of occupying a certain state at each visit. We calculated hazard ratios (HR) and 95%CI to compare SARS-CoV-2 incidence between ethnic groups, using the Dutch origin group as a reference, adjusting for current age in years and sex. We specified calendar time as two piecewise-constant functions (January 1-June 30, 2020, i.e. the first wave in the Netherlands; July 1, 2020-March 31, 2021; i.e. the second wave) assuming that the incidence rate would be constant within the epidemic waves of SARS-CoV-2 in the Netherlands.^16^ Cumulative incidence until March 31, 2021 was directly obtained from this model. We additionally tested for interaction between ethnicity and calendar time. In sensitivity analyses, we first examined the effect of using August 15, 2020 as the date demarcating the first and second wave when defining the two piecewise-constant functions. Second, we examined the effect of differential loss to follow-up (LTFU) by including an additional absorbing state for participants who tested SARS-CoV-2 antibody negative at the first visit and did not return for the second visit, for whom follow-up was censored at March 31, 2021.

To identify determinants of SARS-CoV-2 incidence within ethnic groups, we calculated univariable and multivariable HRs and their 95%CI comparing levels of factors using the piecewise-constant functions for calendar time described above. Covariates obtained from both visits were included as time-updated. *P-*values were calculated from the 95%CI of the HR. We constructed a multivariable model by including all covariates for which the variable (for continuous variables) or at least one category (for categorical variables) was associated with *P*<0·2 in univariable analyses. Backward selection was then employed to obtain a final model by sequentially removing variables that were no longer associated (*P*≥0·05). We additionally tested for interaction between each determinant in the final model and calendar time. We furthermore described the distribution of each identified determinant per visit.

Significance was defined at a *P*<0·05. All analyses were conducted using Stata 15·1 (StataCorp, College Station, TX, USA) and the *msm* package^17^ in R version 3·5·2 (Vienna, Austria).

## Results

### Study population

We included 2,497 individuals after the first wave. 503 (20*·*1%) were of Dutch origin, 453 (18.1%) South-Asian Surinamese, 407 (16*·*3%) African Surinamese, 331 (13*·*3%) Ghanaian, 409 (16*·*4%) Turkish, and 394 (15*·*8%) Moroccan. Participant characteristics have been described in detail previously.^12^ Briefly, median age was 54 years (interquartile range [IQR]: 44-61) and 56*·*6% were female. By March 31, 2021, 2,075 (83*·*1%) participants returned for the second visit. The percentage returning was highest for participants of Dutch origin (n=468/503, 93*·*0%), followed by participants of African Surinamese (n=362/407, 88.9%), South-Asian Surinamese (n=394/453, 87*·*0%), Turkish (n=326/409, 79*·*7%), Moroccan (n=312/394, 79*·*2%), and Ghanaian (n=213/331, 64*·*4%) origin. Number of individuals with a visit and proportion testing positive and negative per ethnic group are given for each calendar month in **Supplementary Figure S1**. The age and sex distribution of participants who returned by March 31, 2021 were comparable to participants who did not; however, participants who did not return were more likely to be first generation migrants, not be employed, and have a lower educational and occupational level, difficulties with the Dutch language, lower health literacy, and SARS-CoV-2 antibodies at the first visit (**Table 1**).

**Table 1.** Characteristics of the HELIUS participants included in the COVID-19 seroprevalence substudy, Amsterdam, the Netherlands, June 24, 2020 – March 31, 2021.

| Characteristic | Total<br>(N=2,497) | Participated in<br>round 1 and 2<br>before or on 31<br>March 2021<br>(n=2,075) | Participated in round<br>1 and lost to<br>follow-up in round 2 /<br>second visit after<br>31 March 2021<br>(n=422) | P-value |
| --- | --- | --- | --- | --- |
|  | n (%) | n (%) | n (%) |  |
| **Sex** |  |  |  | 0.067 |
| Male | 1,083 (43.4%) | 917 (44.2%) | 166 (39.3%) |  |
| Female | 1,414 (56.6%) | 1,158 (55.8%) | 256 (60.7%) |  |
| **Age in years on 1 January 2020** |  |  |  | 0.066 |
| Median [IQR] | 54 [44-61] | 54 [44-61] | 53 [43-60] |  |
| **Ethnicity** |  |  |  | <0.001 |
| Dutch | 503 (20.1%) | 468 (22.6%) | 35 (8.3%) |  |
| South-Asian Surinamese | 453 (18.1%) | 394 (19.0%) | 59 (14.0%) |  |
| African Surinamese | 407 (16.3%) | 362 (17.4%) | 45 (10.7%) |  |
| Ghanaian | 331 (13.3%) | 213 (10.3%) | 118 (28.0%) |  |
| Turkish | 409 (16.4%) | 326 (15.7%) | 83 (19.7%) |  |
| Moroccan | 394 (15.8%) | 312 (15.0%) | 82 (19.4%) |  |
| **Migration generation** |  |  |  | <0.001 |
| N/A (Dutch) | 503 (20.1%) | 468 (22.6%) | 35 (8.3%) |  |
| 1<sup>st</sup> | 1,656 (66.3%) | 1,327 (64.0%) | 329 (78.0%) |  |
| 2<sup>nd</sup> | 338 (13.5%) | 280 (13.5%) | 58 (13.7%) |  |
| **City district<sup>a</sup>** |  |  |  | 0.22 |
| Centre | 140 (5.6%) | 117 (5.6%) | 23 (5.5%) |  |
| East | 422 (16.9%) | 363 (17.5%) | 59 (14.0%) |  |
| West | 294 (11.8%) | 239 (11.5%) | 55 (13.0%) |  |
| South | 245 (9.8%) | 214 (10.3%) | 31 (7.3%) |  |
| New-West | 606 (24.3%) | 500 (24.1%) | 106 (25.1%) |  |
| Southeast | 760 (30.4%) | 618 (29.8%) | 142 (33.6%) |  |
| Other/missing | 6 (0.2%) | 5 (0.2%) | 1 (0.2%) |  |
| **Educational level<sup>a</sup>** |  |  |  | <0.001 |
| No school/elementary school | 327 (13.1%) | 240 (11.6%) | 87 (20.6%) |  |
| Lower vocational/lower secondary school | 612 (24.5%) | 499 (24.0%) | 113 (26.8%) |  |
| Intermediary vocational/intermediary secondary school | 700 (28.0%) | 584 (28.1%) | 116 (27.5%) |  |
| Higher vocational/university | 792 (31.7%) | 708 (34.1%) | 84 (19.9%) |  |
| Missing | 66 (2.6%) | 44 (2.1%) | 22 (5.2%) |  |
| **Labor participation<sup>b</sup>** |  |  |  | 0.010 |
| Employed | 1,659 (66.4%) | 1,414 (68.1%) | 245 (58.1%) |  |
| Not in workforce | 309 (12.4%) | 253 (12.2%) | 56 (13.3%) |  |
| Unemployed/on benefits | 300 (12.0%) | 237 (11.4%) | 63 (14.9%) |  |
| Disabled | 151 (6.0%) | 122 (5.9%) | 29 (6.9%) |  |
| Unknown/missing | 26 (1.0%) | 18 (0.9%) | 8 (1.9%) |  |
| **Occupational level<sup>a</sup>** |  |  |  | <0.001 |
| Elementary occupations | 323 (12.9%) | 241 (11.6%) | 82 (19.4%) |  |
| Lower occupations | 537 (21.5%) | 434 (20.9%) | 103 (24.4%) |  |
| Intermediary occupations | 599 (24.0%) | 524 (25.3%) | 75 (17.8%) |  |
| Higher occupations | 500 (20.0%) | 443 (21.3%) | 57 (13.5%) |  |
| Scientific occupations | 202 (8.1%) | 180 (8.7%) | 22 (5.2%) |  |
| Missing | 284 (11.4%) | 222 (10.7%) | 62 (14.7%) |  |
| **Job setting<sup>b</sup>** |  |  |  | 0.036 |
| No job / caretaker only | 741 (29.7%) | 599 (28.9%) | 142 (33.6%) |  |
| Job with no contact within 1.5 meter | 387 (15.5%) | 315 (15.2%) | 72 (17.1%) |  |
| Other job with contact within 1.5 meter | 790 (31.6%) | 663 (32.0%) | 127 (30.1%) |  |
| Child care/schools/higher education | 215 (8.6%) | 194 (9.3%) | 21 (5.0%) |  |
| Bar/restaurant | 69 (2.8%) | 58 (2.8%) | 11 (2.6%) |  |
| Hospital/long-term care facility/Health care worker elsewhere | 288 (11.5%) | 242 (11.7%) | 46 (10.9%) |  |
| Missing | 7 (0.3%) | 4 (0.2%) | 3 (0.7%) |  |
| **Difficulty with Dutch language<sup>a</sup>** |  |  |  | <0.001 |
| N/A (Dutch) | 503 (20.1%) | 468 (22.6%) | 35 (8.3%) |  |
| No | 1,148 (46.0%) | 984 (47.4%) | 164 (38.9%) |  |
| Yes | 782 (31.3%) | 582 (28.0%) | 200 (47.4%) |  |
| Missing | 64 (2.6%) | 41 (2.0%) | 23 (5.5%) |  |
| **Health literacy (SBSQ)<sup>a</sup>** |  |  |  | <0.001 |
| Adequate | 2,164 (86.7%) | 1,843 (88.8%) | 321 (76.1%) |  |
| Low | 274 (11.0%) | 194 (9.3%) | 80 (19.0%) |  |
| Missing | 59 (2.4%) | 38 (1.8%) | 21 (5.0%) |  |
| **Body Mass Index (kg/m<sup>2</sup>), median (IQR)<sup>a</sup>** |  |  |  | <0.001 |
|  | 26 [24-29] | 26 [23-29] | 27 [24-31] |  |
| **SARS-CoV-2 antibody test result round 1<sup>b</sup>** |  |  |  |  |
| Equivocal | 8 (0.3%) | 4 (0.2%) | 4 (0.9%) | <0.001 |
| Negative | 2,250 (90.1%) | 1,896 (91.4%) | 354 (83.9%) |  |
| Positive | 225 (9.0%) | 170 (8.2%) | 55 (13.0%) |  |
| Missing | 14 (0.6%) | 5 (0.2%) | 9 (2.1%) |  |

**Abbreviations:** BMI, body mass index; HELIUS, Healthy Life in an Urban Setting; IQR, interquartile range; N.A., not applicable; SBSQ, Set of Brief Screening Question <sup>a</sup> Presumed higher exposure categories had priority, i.e. if someone was working in a school and as a health care worker, they were categorized as a health care worker. Caretakers were not included as a category because many had other jobs.
<sup>a</sup> Measured at baseline (2011-2015) <sup>b</sup> Measured at COVID-1 visit (2020) <sup>c</sup> Based on self-report, increased fasting glucose ( $\geq 7$ mmol/l) or use of glucose-lowering medication <sup>d</sup> Based on self-report, SBP $\geq 140$ mmHg, DBP $\geq 90$ or blood pressure-lowering medication

### SARS-CoV-2 incidence

At the first visit, 2,483 (99·4%) had a SARS-CoV-2 antibody test result: 225 were positive, 2,250 negative, and 8 had an equivocal result. Of the 2,075 participants returning for the second visit by March 31, 2021, 2,062 (99·4%) had a test result, of whom 3 had been vaccinated, 490 were positive, 1,567 negative, and 2 had an equivocal result. After excluding vaccinated individuals (second visit only) or those with equivocal results, the highest percentage testing positive at the first and second visits, respectively, was observed in the Ghanaian group (95/327, 29·1% and 103/212, 48·6%), followed by Moroccan (32/391, 8·2% and 101/309, 32·7%), Turkish (30/408, 7·4% and 97/321, 30·2%), African Surinamese (22/400, 5·5% and 71/360, 19·7%), South-Asian Surinamese (22/451, 4·9% and 70/391, 17·9%) groups, while the lowest was observed in the Dutch origin group (24/498, 4·8% and 48/416, 11·5%). Among those who returned for the second visit, all 169 participants with SARS-CoV-2 antibodies at the first visit also tested positive at the second visit after a median of 150 days (IQR=138-164).

Estimated cumulative SARS-CoV-2 incidence from January 1, 2020 is presented in **Figure 1**. Compared to participants of Dutch origin (cumulative incidence at March 31, 2021=15·9%), SARS-CoV-2 incidence was higher in participants of South-Asian Surinamese (cumulative incidence=25·0%; adjusted hazard ratio [aHR]=1·66;95%CI=1·16-2·40), African Surinamese (cumulative incidence=28·9%; aHR=1·97;95%CI=1·37-2·83), Ghanaian (cumulative incidence=64·6%; aHR=6·00;95%CI=4·33-8·30), Turkish (cumulative incidence=37·0%; aHR=2·67;95%CI=1·89-3·78), and Moroccan origin (cumulative incidence=41·9%; aHR=3·13;95%CI=2·22-4·42). These differences in incidence compared with the Dutch origin group became wider during the second versus first wave for all ethnic minority groups (all p for interaction<0·05) except the Ghanaian group (**Supplementary Table S1**). Results were similar in the sensitivity analyses defining the two piecewise-constant functions at August 15, 2020 and including LTFU as a separate state (**Supplementary Tables S2 and S3, Figure S2**).

**Figure 1.**
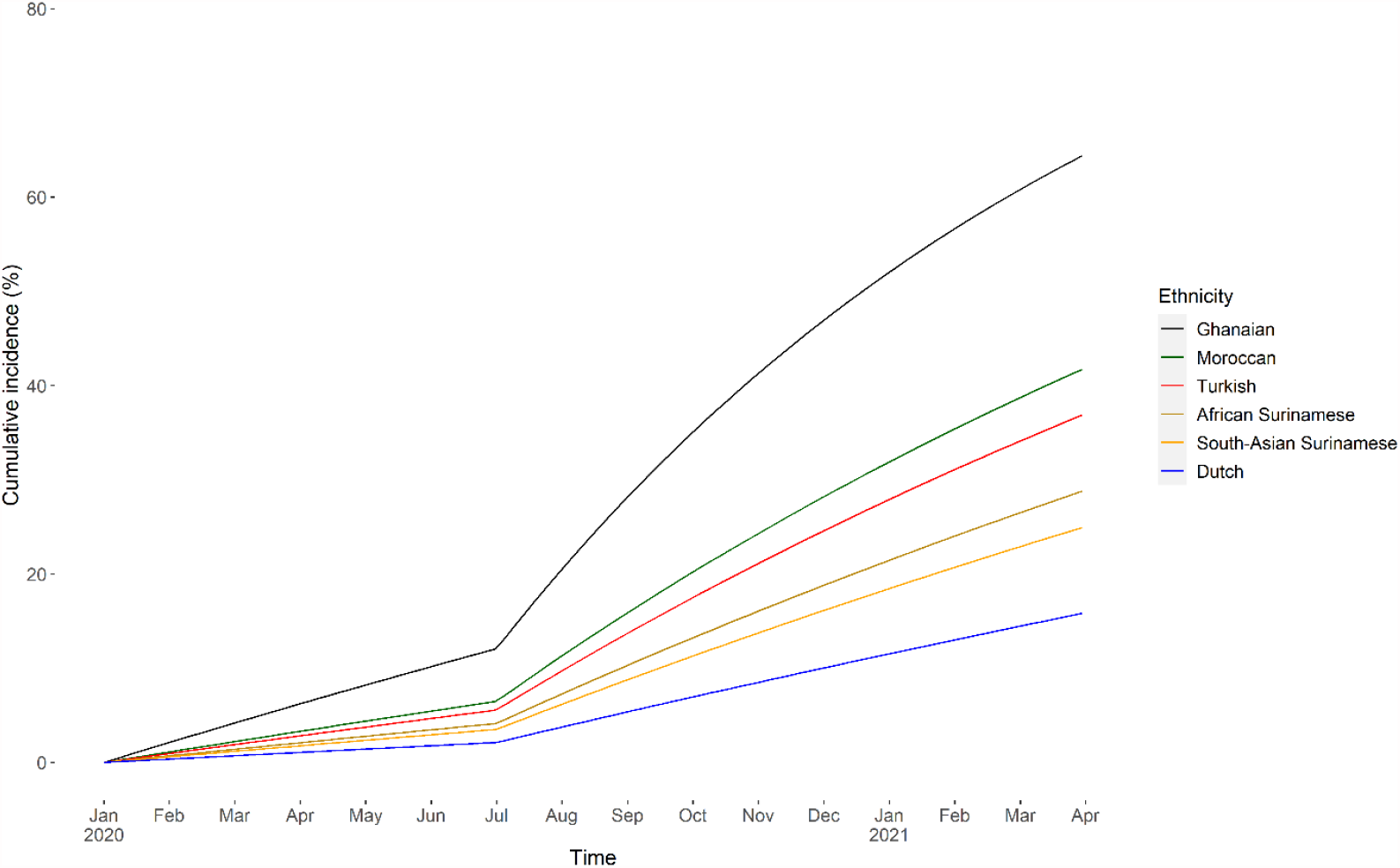
Estimated cumulative SARS-CoV-2 incidence between January 1, 2020 and March 31, 2021 per ethnic group, adjusted for age and sex, HELIUS COVID-19 seroprevalence substudy. Footnote: Incidence was based on SARS-CoV-2 antibody test results from two subsequent study visits. The first visit took place between June 24 and October 9, 2020 and the second between November 23, 2020 and March 31, 2021. We modelled the transition of negative to positive SARS-CoV-2 antibody test using a time-homogenous, continuous-time, two-state Markov model, assuming all participants were SARS-CoV-2 negative on January 1, 2020.

### Determinants of SARS-CoV-2 incidence per ethnic group

Univariable analysis of determinants of SARS-CoV-2 incidence is presented per ethnic group in **Supplementary Tables S4-9**. In multivariable analysis (**Figure 2**), presence of household members suspected of SARS-CoV-2 infection was a determinant of increased incident infection in all groups except the Ghanaian group. Larger household size was associated with increased incident infection in participants of Turkish and Moroccan origin. Low health literacy was associated with increased incident infection in participants of South-Asian Surinamese and African Surinamese origin. Other determinants of increased incident infection were walking or exercising outside in the past week (South-Asian Surinamese), living with an adult child (African Surinamese), having 2 or more visitors in the past week (African Surinamese), walking outside with a dog or kids in the past week (Ghanaian), none/elementary educational level (Turkish), and being a caretaker (Turkish). Determinants associated with decreased incident infection were observed for walking or exercising outside in the past week (African Surinamese), visiting a bar or restaurant in the past week (Ghanaian), living with a child aged 4 through 12 years, doing groceries in the past week and having 1 visitor in the past week (all in Moroccan).

**Figure 2.**
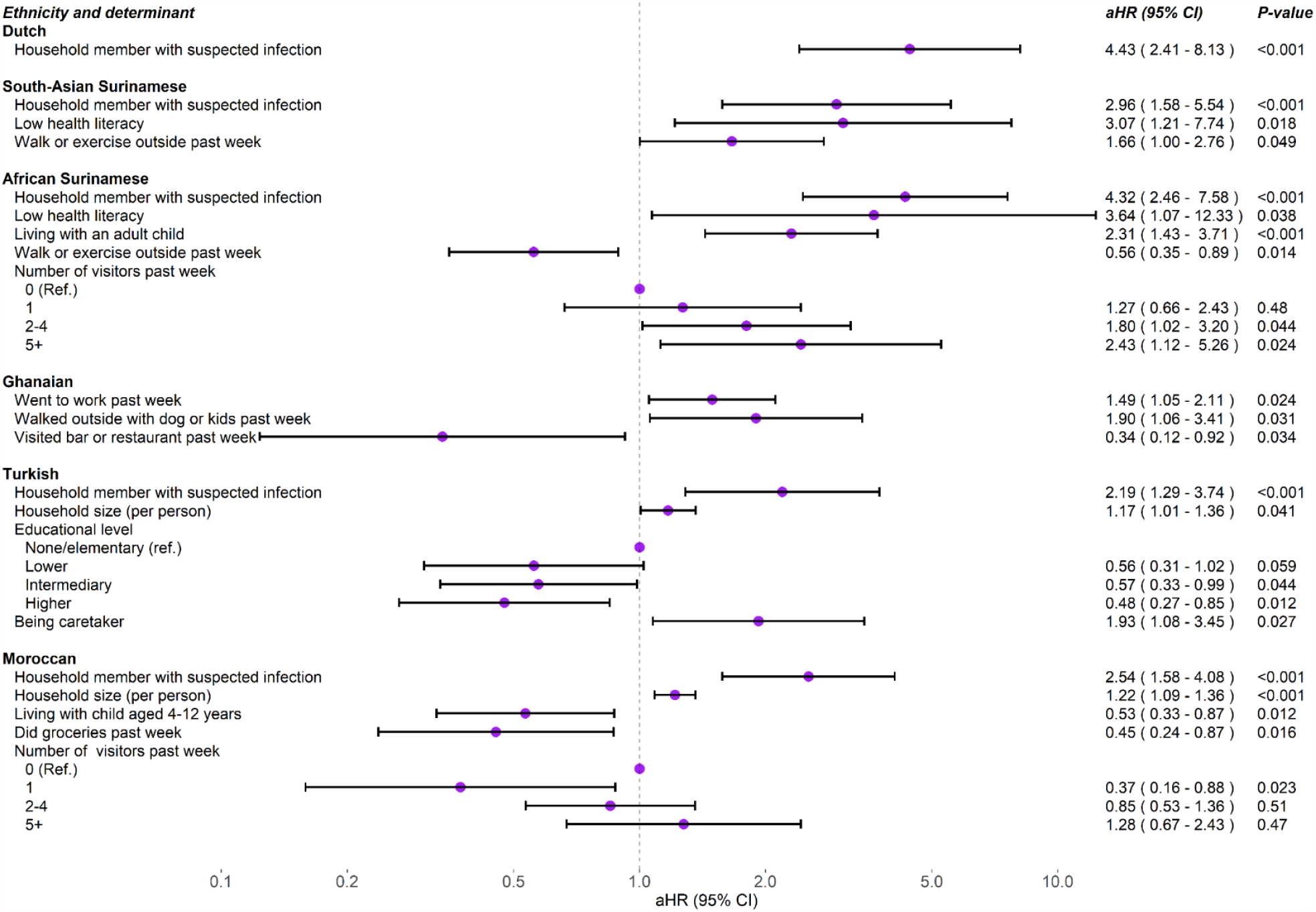
Determinants of SARS-CoV-2 incidence by ethnic group, HELIUS COVID-19 study (multivariable analysis)

The association between presence of household members with suspected SARS-CoV-2 infection and increased incident infection became weaker during the second versus first wave for the Dutch, South-Asian Surinamese, African Surinamese, and Moroccan group (*p* for interaction=0·002, 0·011, 0·004, 0·048, respectively). The association between walking or exercising outside in the past week and increased incident infection in the South-Asian Surinamese group became stronger (*p* for interaction=0·049). Associations with incident infection did not change over calendar time for the other determinants.

The distribution of most time-updated determinants was not different between the first and second visit, except for the number of unique visitors at home in the past week in the African Surinamese group and visiting a bar or restaurant in the past week in the Ghanaian group, both of which were reported to occur less frequently at the second compared to first visit (**Table 2**).

**Table 2.**
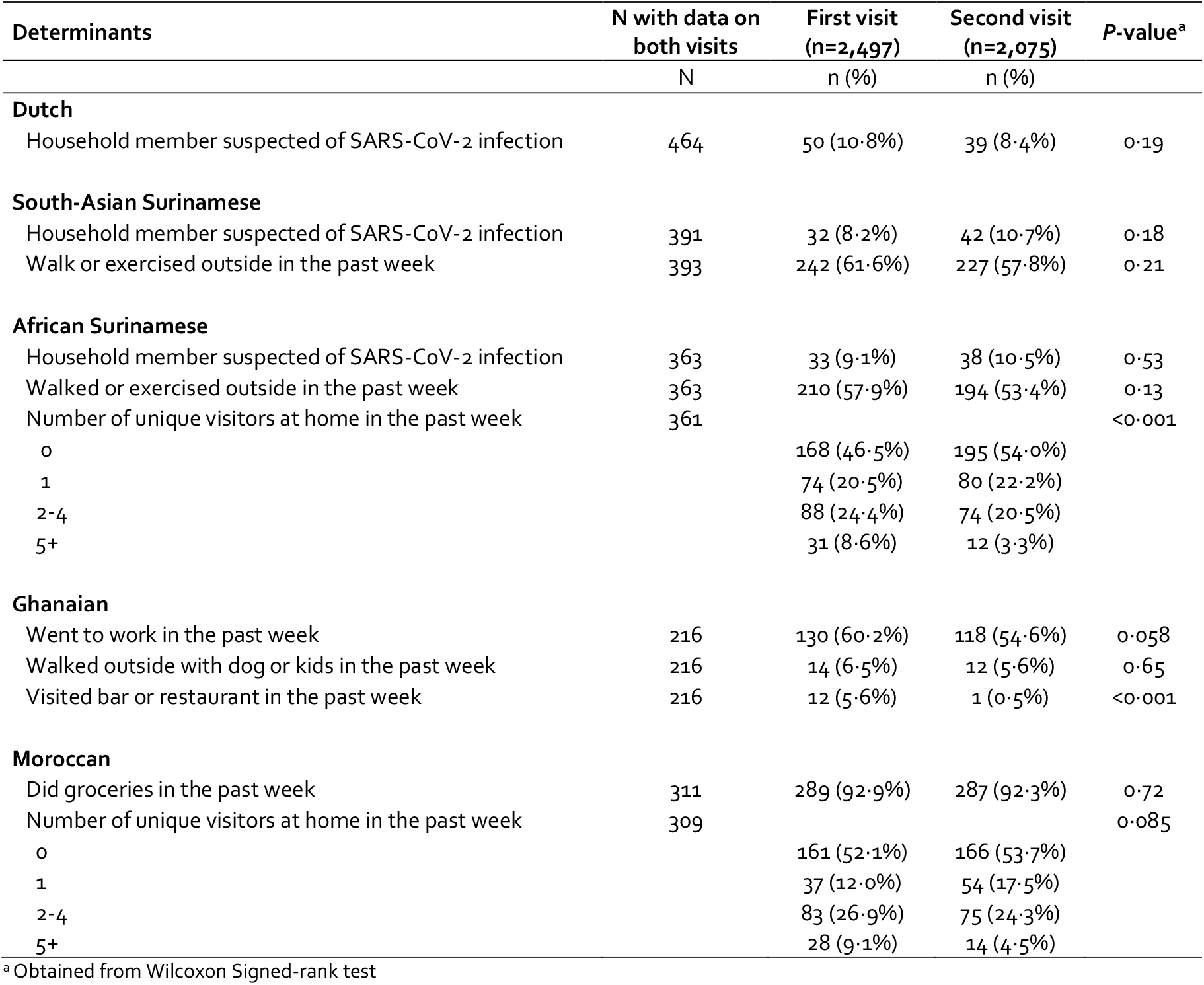
Distribution of time-updated determinants of incident infection per visit, HELIUS COVID-19 seroprevalence substudy, June 24, 2020 – March 31, 2021.

## Discussion

We show that cumulative SARS-CoV-2 incidence until March 31, 2021 was higher in the largest adult ethnic minority groups compared to the adult Dutch origin group in Amsterdam, the Netherlands. These ethnic differences became apparent for all ethnic minority groups during the second wave of the SARS-CoV-2 epidemic, except for individuals of Ghanaian origin, who had the highest incidence from the start of the epidemic onwards. The strongest increases in incidence between the first and second wave were observed in individuals of Turkish and Moroccan origin. Having a household member suspected of SARS-CoV-2 infection, larger household size, and low health literacy were common determinants of SARS-CoV-2 exposure across ethnic groups, whereas some determinants were specific to individual groups. This finding indicates that caution is warranted when broadly generalizing determinants of SARS-CoV-2 incidence.

In the Netherlands, the initial lockdown started shortly (i.e. mid-March) after the first confirmed cases of SARS-CoV-2 in the country on February 27, 2020.^18^ Lockdown measures were gradually lifted from mid-May 2020 onwards, after which the second wave started towards the end of August 2020. The lockdown measures applied after this date until December 2020 were not nearly as restrictive as in the first wave.^19^ During this period, the highest rates of diagnosed SARS-CoV-2 cases per 100,000 inhabitants were observed in the Amsterdam city districts with a relatively lower socioeconomic status and higher number of residents with an ethnic minority background.^20^ This finding suggests that very stringent measures prevented disparities in the initial spread of the virus, but that less stringent measures in the first part of the second wave resulted in a more rapid spread in the largest ethnic minority groups in Amsterdam. An analysis from England, where lockdown measures were stricter than in the Netherlands during the second wave,^19^ corroborates these findings, observing that all ethnic minority groups, except South-Asian groups, less frequently or equally frequently tested positive for SARS-CoV-2 in the second versus first wave, independent of testing uptake.^21^

After the first wave of SARS-CoV-2, targeted prevention efforts towards ethnic minority groups were instated in Amsterdam to reduce disparities in COVID-19. Local prevention teams were deployed to organize outreach activities, such as low-threshold testing in mobile buses/vans circulating in neighborhoods with relatively high number of cases. Our results suggest that these efforts might not have fully prevented further spread and widening of infection rates between ethnic groups. After observing that 26% of the adult Ghanaian group had evidence of past SARS-CoV-2 infection after the first wave, compared to 5-8% in other ethnic minority and Dutch origin groups,^12^ intensified prevention efforts were targeted towards the Ghanaian population. These included discussions of the initial findings with key persons, as well as prevention and information activities in close collaboration with community leaders, general practitioners and employers, online and in common meeting places (i.e. churches, malls). Although these activities might not have had an effect on the increased incidence of Ghanaian individuals of the HELIUS study population, they could have influenced the epidemic in certain settings. For example, we observed that attending religious services in the past week was an important determinant of past SARS-CoV-2 exposure after the first wave in this group,^12^ whereas in the current analysis, this determinant was not retained in the multivariable model. The increased preventive measures during church services could have offered reduced SARS-CoV-2 transmission.

Given the higher rates per population observed in SARS-CoV-2 incidence, as well as hospitalization and mortality in ethnic minority groups,^1,2,22^ which were also apparent in Amsterdam,^4^ sustained and targeted actions to reduce these disparities are warranted. With the wide availability of safe and effective vaccines against SARS-CoV-2, it is imperative to achieve high vaccination uptake, particularly in populations at high risk of SARS-CoV-2 infection and unfavorable outcomes. However, studies predominantly from the USA have demonstrated lower intent to vaccinate against SARS-CoV-2 in ethnic minority groups, although intent varied between groups.^23-25^ In our sample, intent to vaccinate was also lower in all ethnic minority groups compared to those of Dutch origin, especially in the Turkish and Moroccan groups.^26^ Further research should be conducted to identify effective strategies addressing the needs of specific ethnic groups and to promote the uptake of vaccination and other prevention measures.

Having low health literacy was a determinant of increased SARS-CoV-2 incidence in both Surinamese groups and no or elementary education was a determinant in the Turkish group. Although we did not observe any evidence that knowledge of preventive measures was different between ethnic groups from an online survey in a sample of HELIUS participants or that this knowledge differed by educational level within ethnic groups,^27^ low health literacy or educational levels could affect threat appraisal and the ease with which complex behavioral messages could be translated to individual situations, thereby affecting uptake of measures. Targeted provision of comprehensive information on preventive measures in different languages is already ongoing in Amsterdam and these efforts should be continued. In addition, low health literacy and educational level can be proxies for unmeasured behaviors that pose a risk of infection.^28^ As this analysis was exploratory, further research on the exact casual pathways through which ethnicity and socioeconomic factors are implicated in SARS-CoV-2 infection should be conducted.

Household characteristics, such as having a household member suspected of infection and large household size, were common determinants of SARS-CoV-2 incidence across groups. This is in line with previous studies showing that a large part of SARS-CoV-2 transmissions occurs within household settings.^29^ While having a household member suspected of infection did not change over time in our study, this determinant of increased SARS-CoV-2 incidence became weaker within groups during the second wave, suggesting that SARS-CoV-2 transmissions shifted from within the household to other sources, such as public places or workspaces.

Strengths of our study include population-based sampling, with a large number of participants from the major ethnic groups living in Amsterdam, who represented various levels of socioeconomic status and were followed over time. SARS-CoV-2 antibodies were measured twice using a highly sensitive and specific test^30^ at different stages of the epidemic, irrespective of previous COVID-19-related symptoms, which allowed for a less biased estimation of cumulative SARS-CoV-2 incidence. Individual-level determinants of infection were obtained. Nevertheless, there are several limitations. First, our study might be subject to selection bias. Participants in our substudy might have been more concerned about their health compared to non-participants. Second, LTFU differed between ethnic groups, and was higher in participants with lower socio-economic status. We were unable to directly control for differential LTFU, but our sensitivity analysis suggests the impact of LTFU on our results was limited. Third, exposure variables were collected after infection and may have been different at the time of infection, depending on restrictions in place. Fourth, while we included many potential exposures and correlates of infection in our analysis, some might have been missed, such as preventive behaviours (e.g. wearing masks, keeping distance) during activities.

In conclusion, ethnic differences in SARS-CoV-2 infection became apparent during the second wave of infection in the Netherlands and incidence was higher in ethnic minority groups compared to those of Dutch origin. Targeted prevention efforts following the first wave might not have been sufficient to prevent these disparities. Focus should be placed on reaching high vaccine coverage in all ethnic groups, alongside improvement of other targeted prevention strategies addressing the needs of individuals within these groups.

## Data Availability

The HELIUS data are owned by the Amsterdam UMC, location AMC, in Amsterdam, The Netherlands. Any researcher can request the data by submitting a proposal to the HELIUS Executive Board as outlined at http://www.heliusstudy.nl/en/researchers/collaboration, by. The HELIUS Executive Board will check proposals for compatibility with the general objectives, ethical approvals and informed consent forms of the HELIUS study. There are no other restrictions to obtaining the data and all data requests will be processed in the same manner.

## Author contributions

MP, KS, JS and CA conceived, designed, or oversaw the study. HG, AK and JS were involved in the acquisition of data. LC conducted the statistical analysis and drafted the manuscript under supervision of AB and MP. All authors read and approved the final manuscript.

## Acknowledgements

The authors would like to acknowledge the HELIUS COVID-19 study participants for their contribution and the HELIUS team for data collection and management.

## References

1. Sze S, Pan D, Nevill CR, et al. Ethnicity and clinical outcomes in COVID-19: A systematic review and meta-analysis. EClinicalMedicine 2020;29:100630.

2. Public Health England. Disparities in the risk and outcomes of COVID-19. 2020. Available from: https://www.gov.uk/government/publications/covid-19-review-of-disparities-in-risks-and-outcomes. Accessed 20 July 2021.

3. Indseth T, Grosland M, Arnesen T, et al. COVID-19 among immigrants in Norway, notified infections, related hospitalizations and associated mortality: A register-based study. Scand J Public Health 2021;49(1):48–56.

4. Coyer L, Wynberg E, Buster M, et al. Hospitalisation rates differed by city district and ethnicity during the first wave of COVID-19 in Amsterdam, the Netherlands. medRxiv 2021:2021.03.15.21253597.

5. Ward H, Cooke G, Whitaker M, et al. REACT-2 Round 5: increasing prevalence of SARS-CoV- 2 antibodies demonstrate impact of the second wave and of vaccine roll-out in England. medRxiv 2021:2021.02.26.21252512.

6. Hollis ND, Li W, Van Dyke ME, et al. Racial and Ethnic Disparities in Incidence of SARS-CoV- 2 Infection, 22 US States and DC, January 1-October 1, 2020. Emerg Infect Dis 2021;27(5):1477–81.

7. Martin CA, Jenkins DR, Minhas JS, et al. Socio-demographic heterogeneity in the prevalence of COVID-19 during lockdown is associated with ethnicity and household size: Results from an observational cohort study. EClinicalMedicine 2020;25:100466.

8. Bhala N, Curry G, Martineau AR, Agyemang C, Bhopal R. Sharpening the global focus on ethnicity and race in the time of COVID-19. Lancet 2020;395(10238):1673–6.

9. Bambra C, Riordan R, Ford J, Matthews F. The COVID-19 pandemic and health inequalities. J Epidemiol Community Health 2020;74(11):964–8.

10. Webb Hooper M, Napoles AM, Perez-Stable EJ. COVID-19 and Racial/Ethnic Disparities. JAMA 2020;323(24):2466–7.

11. Statistics Netherlands (CBS). Bevolking; leeftijd, migratieachtergrond, geslacht, regio, 1 jan. 1996-2020. Available from: https://opendata.cbs.nl/statline/#/CBS/nl/dataset/37713/table?fromstatweb. Accessed 6 January 2021.

12. Coyer L, Boyd A, Schinkel J, et al. SARS-CoV-2 antibody prevalence and determinants of six ethnic groups living in Amsterdam, the Netherlands: a population-based cross-sectional study, June-October 2020. medRxiv 2021:2021.03.08.21252788.

13. Snijder MB, Galenkamp H, Prins M, et al. Cohort profile: the Healthy Life in an Urban Setting (HELIUS) study in Amsterdam, The Netherlands. BMJ Open 2017;7(12):e017873.

14. GeurtsvanKessel CH, Okba NMA, Igloi Z, et al. An evaluation of COVID-19 serological assays informs future diagnostics and exposure assessment. Nat Commun 2020;11(1):3436.

15. Kalbfleisch JD, Lawless JF. The Analysis of Panel Data under a Markov Assumption. Journal of the American Statistical Association 1985;80(392):863–71.

16. National Institute for Public Health and the Environment (RIVM). Ontwikkeling COVID-19 in grafieken. Available from: https://www.rivm.nl/en/node/154271. Accessed 7 June 2021.

17. Jackson CH. Multi-State Models for Panel Data: The msm Package for R. Journal of Statistical Software 2011;38(8):1–29.

18. Dutch Government (Rijksoverheid). Nieuwe maatregelen tegen verspreiding coronavirus in Nederland. Available from: https://www.rijksoverheid.nl/actueel/nieuws/2020/03/12/nieuwe-maatregelen-tegen-verspreiding-coronavirus-in-nederland. Accessed 7 June 2012.

19. Hale T, Angrist N, Goldszmidt R, et al. A global panel database of pandemic policies (Oxford COVID-19 Government Response Tracker). Nat Hum Behav 2021;5(4):529–38.

20. Public Health Service of Amsterdam. Corona in cijfers in Amsterdam-Amstelland. Available from: https://www.ggd.amsterdam.nl/coronavirus/situatie-regio-amsterdam-amstelland/. Accessed 16 June 2021.

21. Mathur R, Rentsch CT, Morton CE, et al. Ethnic differences in SARS-CoV-2 infection and COVID-19-related hospitalisation, intensive care unit admission, and death in 17 million adults in England: an observational cohort study using the OpenSAFELY platform. Lancet 2021;397(10286):1711–24.

22. Raharja A, Tamara A, Kok LT. Association Between Ethnicity and Severe COVID-19 Disease: a Systematic Review and Meta-analysis. J Racial Ethn Health Disparities 2020:1–10.

23. Robinson E, Jones A, Lesser I, Daly M. International estimates of intended uptake and refusal of COVID-19 vaccines: A rapid systematic review and meta-analysis of large nationally representative samples. Vaccine 2021;39(15):2024–34.

24. Malik AA, McFadden SM, Elharake J, Omer SB. Determinants of COVID-19 vaccine acceptance in the US. EClinicalMedicine 2020;26:100495.

25. Razai MS, Osama T, McKechnie DGJ, Majeed A. Covid-19 vaccine hesitancy among ethnic minority groups. BMJ 2021;372:513.

26. Public Health Service of Amsterdam. Corona en etniciteit. 2021. Available from: https://www.ggd.amsterdam.nl/nieuwsoverzicht/amsterdammers-migratieachtergrond/. Accessed 20 June 2021.

27. Stronks K, Prins M, Agyemang C. Bevolkingsgroepen met een migratieachtergrond zwaarder getroffen door COVID-19. 2021. Available from: https://www.ggd.amsterdam.nl/nieuwsoverzicht/amsterdammers-migratieachtergrond/. Accessed 20 June 2021.

28. Paakkari L, Okan O. COVID-19: health literacy is an underestimated problem. Lancet Public Health 2020;5(5):e249–e50.

29. Sette A, Crotty SA-O. Pre-existing immunity to SARS-CoV-2: the knowns and unknowns. Nat Rev Immunol 2020;20(8):457–8.

30. Zonneveld R, Jurriaans S, van Gool T, Hofstra JJ, Hekker TAM, Defoer P, et al. Head-to-head validation of six immunoassays for SARS-CoV-2 in hospitalized patients. J Clin Virol 2021;139:104821.

