## Appendix for "Ethnic disparities in incident SARS-CoV-2 infections became wider during the second wave of SARS-CoV-2 in Amsterdam, the Netherlands: a population-based longitudinal study"

| **Contents** | **Page** |
|---|---|
| Figure S1. Number of individuals with a visit and test results per month by ethnicity, Amsterdam, the Netherlands, June 24, 2020 – May 7, 2021 | 2 |
| Table S1. Comparison of SARS-CoV-2 incidence since 1 January 2020 between ethnic groups, including interaction terms between ethnicity and time period before/after July 1, 2020. | 3 |
| Table S2. Estimated cumulative SARS-CoV-2 incidence between January 1, 2020 and March 31, 2021 in sensitivity analyses compared to main analysis. | 4 |
| Table S3. Interaction terms between ethnicity and time period in sensitivity analyses compared to main analysis. | 4 |
| Figure S2. Estimated cumulative SARS-CoV-2 incidence between January 1, 2020 and March 31, 2021 per ethnic group, adjusted for age and sex, defining the two piecewise-constant functions on August 15, 2021, HELIUS COVID-19 seroprevalence substudy | 5 |
| Table S4. Factors associated with transitioning to positive SARS-CoV-2 antibody status in Dutch participants, Amsterdam, the Netherlands, January 1, 2020 – March 31, 2021 | 6 |
| Table S5. Factors associated with transitioning to positive SARS-CoV-2 antibody status in South-Asian Surinamese participants, Amsterdam, the Netherlands, January 1, 2020 – March 31, 2021 | 8 |
| Table S6. Factors associated with transitioning to positive SARS-CoV-2 antibody status in African Surinamese participants, Amsterdam, the Netherlands, January 1, 2020 – March 31, 2021 | 10 |
| Table S7. Factors associated with transitioning to positive SARS-CoV-2 antibody status in Ghanaian participants, Amsterdam, the Netherlands, January 1, 2020 – March 31, 2021 | 12 |
| Table S8. Factors associated with transitioning to positive SARS-CoV-2 antibody status in Turkish participants, Amsterdam, the Netherlands, January 1, 2020 – March 31, 2021 | 14 |
| Table S9. Factors associated with transitioning to positive SARS-CoV-2 antibody status in Moroccan participants, Amsterdam, the Netherlands, January 1, 2020 – March 31, 2021 | 16 |

**Figure S1. Number of individuals with a visit and test results per month by ethnicity, Amsterdam, the Netherlands, June 24, 2020 – May 7, 2021**

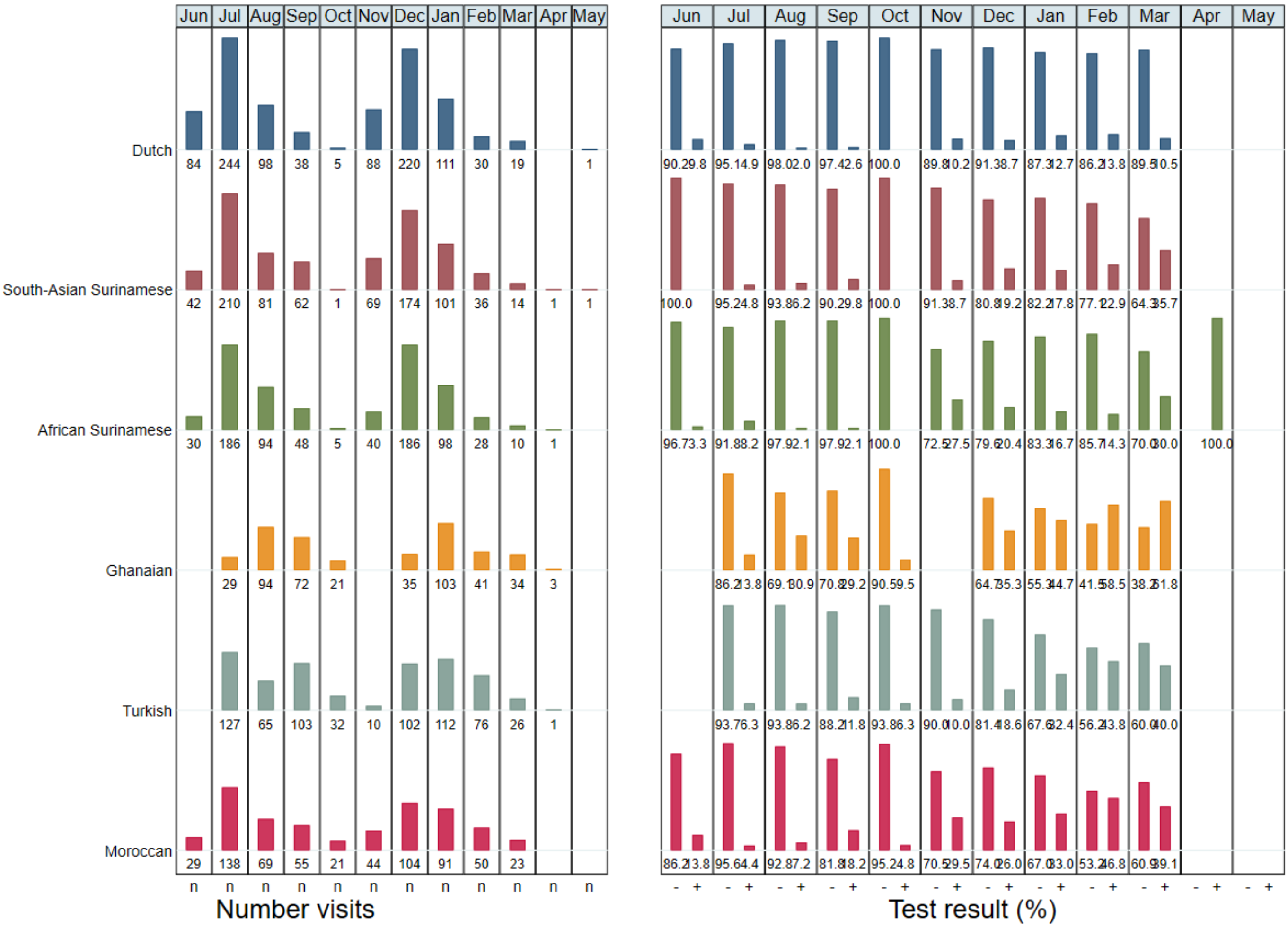


The left side of the graph shows the number of individuals with a visit in the COVID-19 seroprevalence substudy, per calendar month by ethnic group. The right side of the graph shows the distribution of test results per month by ethnic group, excluding people without a test result, equivocal test result or who were already vaccinated. The first visit took place between June 24 and October 9, 2020 and the second visit between November 23, 2020 and June 4, 2021. The cut-off date for this figure was on May 7, 2021.

**Table S1. Comparison of SARS-CoV-2 incidence since 1 January 2020 between ethnic groups, including interaction terms between ethnicity and time period before/after July 1, 2020.**

| Characteristic | aHR (95% CI) | P-value |
|---|---|---|
| **Ethnicity** | | |
| Dutch | 1 | |
| South-Asian Surinamese | 0·58 (0·24-1·40) | 0·23 |
| African Surinamese | 0·93 (0·46-1·90) | 0·86 |
| Ghanaian | 4·74 (2·66-8·45) | <0·001 |
| Turkish | 0·42 (0·12-1·56) | 0·20 |
| Moroccan | 0·78 (0·34-1·79) | 0·56 |
| **Sex** | | |
| Male | 1 | |
| Female | 1·12 (0·94-1·33) | 0·19 |
| **Per year increase in age in years** | 1·00 (0·99-1·01) | 0·53 |
| **Time period** | | |
| Before 1 July 2021 | 1 | |
| On or after 1 July 2021 | 1·57 (0·83-2·96) | 0·17 |
| **Interaction term ethnicity*time period** | | |
| Dutch on or after 1 July 2021 | 1 | |
| South-Asian Surinamese on or after 1 July 2021 | 4·30 (1·50-12·3) | 0·007 |
| African Surinamese on or after 1 July 2021 | 2·98 (1·22-7·31) | 0·017 |
| Ghanaian on or after 1 July 2021 | 1·39 (0·61-3·20) | 0·44 |
| Turkish on or after 1 July 2021 | 10·6 (2·50-44·6) | 0·001 |
| Moroccan on or after 1 July 2021 | 6·52 (2·43-17·5) | <0·001 |

**Abbreviations:** aHR, adjusted hazard ratio; CI, confidence interval

**Table S2. Estimated cumulative SARS-CoV-2 incidence between January 1, 2020 and March 31, 2021 in sensitivity analyses compared to main analysis.**

| | Main analysis: piecewise-constant based on July 1, 2020 | | | Sensitivity analysis 1: piecewise-constant based on August 15, 2020 | | | Sensitivity analysis 2: LTFU Included as absorbing state | | |
|---|---|---|---|---|---|---|---|---|---|
| | **Cumulative incidence (%)** | **aHR (95% CI)** | **P-value** | **Cumulative incidence (%)** | **aHR (95% CI)** | **P-value** | **Cumulative incidence (%)** | **aHR (95% CI)** | **P-value** |
| **Ethnicity** | | | | | | | | | |
| Dutch | 15·9 (9·6-25·5) | 1 | | 16·7 (10·3-26·3) | 1 | | 14·4 (0-22·2) | 1 | |
| South-Asian Surinamese | 25·0 (15·8-37·9) | 1·66 (1·16-2·40) | 0·006 | 26·3 (17·4-39·4) | 1·67 (1·16-2·41) | 0·006 | 21·3 (0-31·5) | 1·62 (1·12-2·33) | 0·010 |
| African Surinamese | 28·9 (18·8-43·6) | 1·97 (1·37-2·83) | <0·001 | 30·3 (18·7-46·0) | 1·98 (1·38-2·83) | <0·001 | 25·5 (0-36·9) | 1·94 (1·35-2·79) | <0·001 |
| Ghanaian | 64·6 (49·6-80·1) | 6·00 (4·33-8·30) | <0·001 | 67·7 (50·9-83·5) | 6·18 (4·46-8·56) | <0·001 | 50·8 (0-63·8) | 5·47 (3·95-7·58) | <0·001 |
| Turkish | 37·0 (25·3-50·0) | 2·67 (1·89-3·78) | <0·001 | 38·8 (27·6-53·8) | 2·69 (1·90-3·80) | <0·001 | 29·9 (0-42·1) | 2·52 (1·78-3·56) | <0·001 |
| Moroccan | 41·9 (30·0-58·1) | 3·13 (2·22-4·42) | <0·001 | 43·6 (30·4-60·0) | 3·14 (2·23-4·42) | <0·001 | 33·7 (0-45·3) | 2·93 (2·08-4·13) | <0·001 |

**Abbreviations:** aHR, adjusted hazard ratio; CI, confidence interval; LTFU, loss to follow-up

**Table S3. Interaction terms between ethnicity and time period in sensitivity analyses compared to main analysis.**

| | Main | Sensitivity 1 | Sensitivity 2 |
|---|---|---|---|
| | **P-value** | **P-value** | **P-value** |
| **Interaction term ethnicity*time period** | | | |
| Dutch | Ref | Ref | Ref |
| South-Asian Surinamese | 0·007 | 0·012 | 0·007 |
| African Surinamese | 0·017 | 0·017 | 0·017 |
| Ghanaian | 0·44 | 0·99 | 0·44 |
| Turkish | 0·001 | <0·001 | 0·001 |
| Moroccan | <0·001 | <0·001 | <0·001 |

**Abbreviations:** aHR, adjusted hazard ratio; CI, confidence interval

**Figure S2. Estimated cumulative SARS-CoV-2 incidence between January 1, 2020 and March 31, 2021 per ethnic group, adjusted for age and sex, defining the two piecewise-constant functions on August 15, 2021, HELIUS COVID-19 seroprevalence substudy**

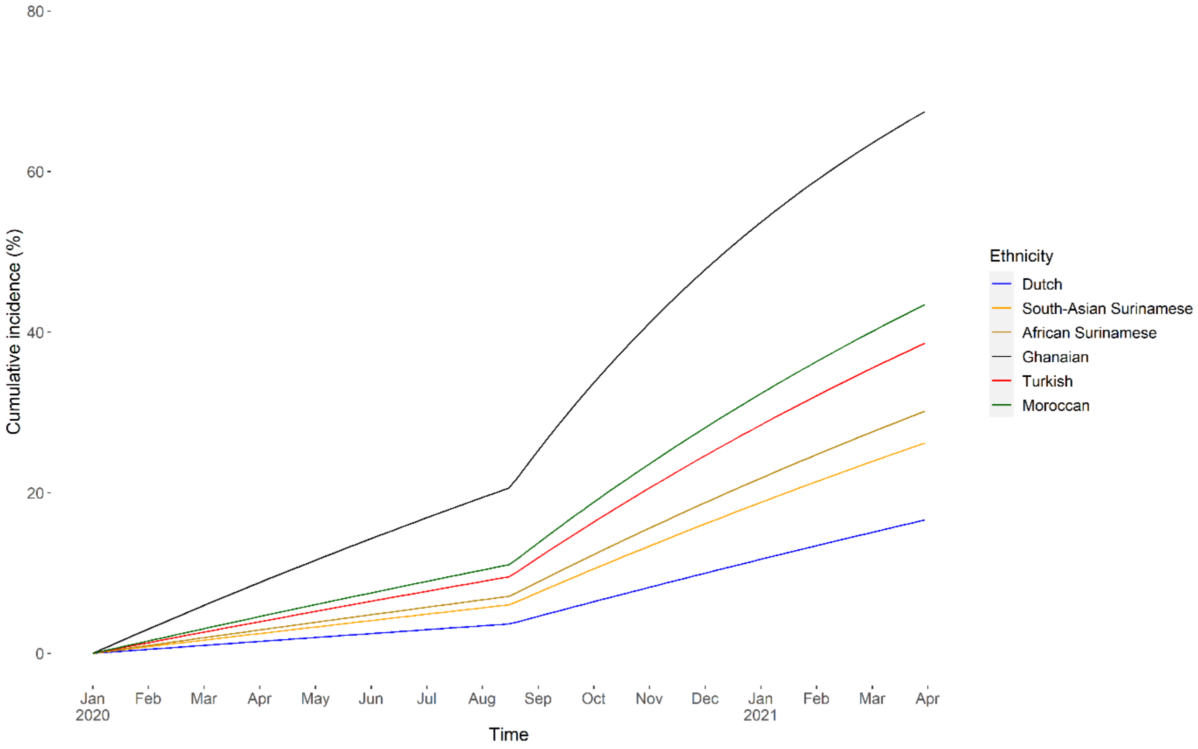


Footnote: Incidence was based on SARS-CoV-2 antibody test results from two subsequent study visits. The first visit took place between June 24 and October 9, 2020 and the second between November 23, 2020 and March 31, 2021. We chose to administratively censor follow-up on March 31, 2021 given the few participants with testing after this date (see also **Fig S1**). We modelled the transition of negative to positive SARS-CoV-2 antibody test using a time-homogenous, continuous-time, two-state Markov model, assuming all participants were SARS-CoV-2 negative on January 1, 2020.

**Table S4. Factors associated with transitioning to positive SARS-CoV-2 antibody status in Dutch participants, Amsterdam, the Netherlands, January 1, 2020 – March 31, 2021**

| | Univariable | | Multivariable | |
|---|---|---|---|---|
| **Characteristic** | **HR (95% CI)** | **P-value** | **aHR (95% CI)** | **P-value** |
| **Sex** | | | | |
| Male | 1 | | | |
| Female | 1·37 (0·77-2·44) | 0·28 | | |
| **Per year increase in age in years** | 1·00 (0·98-1·02) | 0·87 | | |
| **Migration generation** | | | | |
| 1st | NA | | | |
| 2nd | NA | | | |
| **Educational level** [c] | | | | |
| No school/elementary school | 1 | | | |
| Lower vocational/ lower secondary school | 0·18 (0·02-1·25) | 0·082 | | |
| Intermediary vocational/ intermediary secondary school | 0·40 (0·09-1·89) | 0·25 | | |
| Higher vocational/university | 0·56 (0·35-2·33) | 0·44 | | |
| **Labor participation** [c] | | | | |
| Employed | 1 | | | |
| Not in workforce | 0·77 (0·34-1·72) | 0·70 | | |
| Unemployed/on benefits | 0·88 (0·21-3·62) | 0·83 | | |
| Disabled | 0·86 (0·17-6·25) | 0·81 | | |
| Unknown/missing | 0 (0-inf) | 0·95 | | |
| **Elementary occupation** [c] | | | | |
| No | 1 | | | |
| Yes | 1·12 (0·64-1·94) | 0·71 | | |
| **Difficulties with Dutch language** | | | | |
| No | NA | | | |
| Yes | NA | | | |
| **Health literacy (SBSQ)** [c] | | | | |
| Adequate | 1 | | | |
| Low | 0 (0-inf) | 0·91 | | |
| **Job setting**[b,e] | | | | |
| No job / caretaker only | 1 | | | |
| Job with no contact within 1·5 meter | 0·95 (0·36-2·56) | 0·93 | | |
| Other job with contact within 1·5 meter | 1·54 (0·69-3·46) | 0·30 | | |
| Child care/schools/higher education | 1·86 (0·74-4·68) | 0·19 | | |
| Bar/restaurant | 1·12 (1·42-8·85) | 0·92 | | |
| Hospital/long-term care facility/Care worker elsewhere | 1,14 (0·41-3·21) | 0·81 | | |
| **Caretaker** [b] | | | | |
| No | 1 | | | |
| Yes | 1·31 (0·64-2·70) | 0·47 | | |
| **Per 1 person increase in number of household members**[c] | 1·25 (0·98-1·58) | 0·068 | | |
| **Lives with other people** [b] | | | | |
| **Partner** | 1·73 (0·89-3·39) | 0·11 | | |
| **Children up to 3 years old** | 0·51 (0·12-2·09) | 0·35 | | |
| **Children 4 through 12 years old** | 0·87 (0·34-2·19) | 0·78 | | |
| **Children 13 through 17 years old** | 1·26 (0·45-3·51) | 0·67 | | |
| **Children 18+ years old** | 1·51 (0·68-3·36) | 0·32 | | |
| **Parents or parents-in-law** | 0 (0-inf) | 0·91 | | |
| **Other adults** | 0·53 (0·07-3·82) | 0·54 | | |

| | | | | |
|---|---|---|---|---|
| **Household member/steady partner with suspected infection[b]** | | | | |
| N·A·/No | 1 | | 1 | |
| Yes | 4·43 (2·41-8·13) | <0·001 | 4·43 (2·41-8·13) | <0·001 |
| **Number of times left home in the past week [b,d] (per 1 increase)** | 1·02 (0·99-1·05) | 0·28 | | |
| 0-7 | 1 | | | |
| 8-11 | 1·68 (0·58-4·84) | 0·34 | | |
| 12-16 | 0·60 (0·19-1·89) | 0·39 | | |
| 17+ | 1·43 (0·55-3·73) | 0·47 | | |
| **In the past week, left home to [b]:** | | | | |
| **Work** | 1·62 (0·92-2·86) | 0·097 | | |
| **Do groceries** | 0·78 (0·24-2·52) | 0·70 | | |
| **Visit family or friends** | 1·16 (0·63-2·14) | 0·64 | | |
| **Walk the dog or go outside with kids** | 1·08 (0·57-2·04) | 0·82 | | |
| **Walk or exercise outside** | 0·70 (0·37-1·32) | 0·28 | | |
| **Take care of someone** | 1·30 (0·63-2·68) | 0·48 | | |
| **Pick up prescription medicines or visit doctor** | 1·73 (0·93-3·22) | 0·082 | | |
| **Attend religious service** | 2·16 (0·3-15·67) | 0·46 | | |
| **Visit cultural place** | 1·00 (0·47-2·12) | 0·99 | | |
| **Visit bar or restaurant** | 0·80 (0·46-1·40) | 0·44 | | |
| **Indoor sports** | 0·97 (0·41-2·27) | 0·94 | | |
| **Visit recreational park** | 0·80 (0·45-1·43) | 0·46 | | |
| **Frequency of using public transportation in the past week [b]** | | | | |
| 0 days | 1 | | | |
| 1-2 days | 1·27 (0·67-2·41) | 0·48 | | |
| 3-4 days | 1·62 (0·62-4·19) | 0·33 | | |
| 5-7 days | 2·26 (0·54-9·49) | 0·27 | | |
| **Number of unique visitors at home in the past week [b] (per 1 increase)** | 1·03 (0·95-1·12) | 0·50 | | |
| 0 | 1 | | | |
| 1 | 1·35 (0·62-2·93) | 0·45 | | |
| 2-4 | 1·09 (0·53-2·22) | 0·83 | | |
| 5+ | 2·11 (0·92-4·86) | 0·078 | | |
| **Travelled abroad in 2020 [b]** | | | | |
| No | 1 | | | |
| Yes | 1·12 (0·64-1·97) | 0·69 | | |

**Abbreviations:** CI, confidence interval; HELIUS, Healthy Life in an Urban Setting; N.A., not applicable; OR, odds ratio

[a] Those with an equivocal test result were excluded from this analysis [b] Measured at COVID-1 visit (2020) [c] Measured at baseline (2011-2015) [d] Quartiles [e] Presumed higher exposure categories had priority, i.e. if someone was working in a school and as a care worker, they were categorized as a health worker. Caretakers were not included as a category because many had other jobs.

In multivariable analysis, the following variables were removed as they were no longer significant in the multivariable model: number of household members, job setting, living with a partner, leaving home to work in the past week, educational level, number of unique visitors at home in the past week, and leaving home to pick up medication in the past week.

**Table S5. Factors associated with transitioning to positive SARS-CoV-2 antibody status in South-Asian Surinamese participants, Amsterdam, the Netherlands, January 1, 2020 – March 31, 2021**

| | Univariable | | Multivariable | |
|---|---|---|---|---|
| **Characteristic** | **HR (95% CI)** | **P-value** | **aHR (95% CI)** | **P-value** |
| **Sex** | | | | |
| Male | 1 | | | |
| Female | 1·32 (0·80-2·17) | 0·28 | | |
| **Per year increase in age in years** | 1·00 (0·98-1·02) | 0·74 | | |
| **Migration generation** | | | | |
| 1st | 1 | | | |
| 2nd | 1·44 (0·83-2·48) | 0·20 | | |
| **Educational level** [c] | | | | |
| No school/elementary school | 1 | | | |
| Lower vocational/ lower secondary school | 1·14 (0·54-2·41) | 0·75 | | |
| Intermediary vocational/ intermediary secondary school | 0·90 (0·41-1·98) | 0·81 | | |
| Higher vocational/university | 0·78 (0·33-1·81) | 0·57 | | |
| **Labor participation** [c] | | | | |
| Employed | 1 | | | |
| Not in workforce | 0·62 (0·25-1·55) | 0·31 | | |
| Unemployed/on benefits | 0·72 (0·33-1·57) | 0·41 | | |
| Disabled | 0·55 (0·20-1·53) | 0·26 | | |
| Unknown/missing | 1·20 (0·17-8·71) | 0·87 | | |
| **Elementary occupation** [c] | | | | |
| No | 1 | | | |
| Yes | 0·95 (0·66-1·36) | 0·78 | | |
| **Difficulties with Dutch language** | | | | |
| No | 1 | | | |
| Yes | 1·25 (0·74-2·12) | 0·41 | | |
| **Health literacy (SBSQ)** [c] | | | | |
| Adequate | 1 | | 1 | |
| Low | 2·49 (1·00-6·21) | 0·049 | 3·07 (1·21-7·74) | 0·018 |
| **Job setting**[b,e] | | | | |
| No job / caretaker only | 1 | | | |
| Job with no contact within 1·5 meter | 0·76 (0·32-1·79) | 0·54 | | |
| Other job with contact within 1·5 meter | 1·26 (0·71-2·23) | 0·43 | | |
| Child care/schools/higher education | 2·06 (0·91-4·65) | 0·083 | | |
| Bar/restaurant | 1·33 (0·31-5·66) | 0·72 | | |
| Hospital/long-term care facility/Care worker elsewhere | 0·81 (0·33-2·00) | 0·66 | | |
| **Caretaker** [b] | | | | |
| No | 1 | | | |
| Yes | 0·90 (0·46-1·76) | 0·78 | | |
| **Per 1 person increase in number of household members**[c] | 1·16 (0·98-1·37) | 0·092 | | |
| **Lives with other people** [b] | | | | |
| **Partner** | 1·34 (0·84-2·15) | 0·22 | | |
| **Children up to 3 years old** | 1·98 (0·79-4·92) | 0·14 | | |
| **Children 4 through 12 years old** | 1·01 (0·48-2·10) | 0·99 | | |
| **Children 13 through 17 years old** | 1·21 (0·56-2·63) | 0·64 | | |
| **Children 18+ years old** | 1·05 (0·64-1·72) | 0·85 | | |
| **Parents or parents-in-law** | 1·93 (0·96-3·90) | 0·065 | | |
| **Other adults** | 0·90 (0·36-2·24) | 0·83 | | |
| **Household member/steady partner with suspected infection**[b] | | | | |

| | | | | |
|---|---|---|---|---|
| N·A·/No | 1 | | 1 | |
| Yes | 2·66 (1·43-4·96) | 0·002 | 2·96 (1·58-5·54) | <0·001 |
| **Number of times left home in the past week [b,d] (per 1 increase)** | 1·02 (0·98-1·05) | 0·30 | | |
| 0-7 | 1 | | | |
| 8-11 | 1·12 (0·59-2·11) | 0·74 | | |
| 12-16 | 1·39 (0·73-2·62) | 0·32 | | |
| 17+ | 1·52 (0·76-3·04) | 0·24 | | |
| **In the past week, left home to [b]:** | | | | |
| **Work** | 0·88 (0·55-1·42) | 0·62 | | |
| **Do groceries** | 1·12 (0·41-3·06) | 0·84 | | |
| **Visit family or friends** | 1·02 (0·64-1·63) | 0·94 | | |
| **Walk the dog or go outside with kids** | 1·79 (0·89-3·60) | 0·10 | | |
| **Walk or exercise outside** | 1·54 (0·93-2·55) | 0·092 | 1·66 (1·00-2·76) | 0·049 |
| **Take care of someone** | 0·73 (0·35-1·52) | 0·40 | | |
| **Pick up prescription medicines or visit doctor** | 0·76 (0·43-1·32) | 0·34 | | |
| **Attend religious service** | 0·95 (0·30-3·02) | 0·93 | | |
| **Visit cultural place** | 1·59 (0·58-4·36) | 0·38 | | |
| **Visit bar or restaurant** | 0·82 (0·44-1·52) | 0·54 | | |
| **Indoor sports** | 1·39 (0·78-2·46) | 0·26 | | |
| **Visit recreational park** | 0·85 (0·44-1·66) | 0·65 | | |
| **Frequency of using public transportation in the past week [b]** | | | | |
| 0 days | 1 | | | |
| 1-2 days | 1·25 (0·69-2·27) | 0·47 | | |
| 3-4 days | 1·44 (0·61-3·36) | 0·41 | | |
| 5-7 days | 1·06 (0·38-2·95) | 0·92 | | |
| **Number of unique visitors at home in the past week [b]** | 1·00 (0·95-1·05) | 0·99 | | |
| 0 | 1 | | | |
| 1 | 0·74 (0·37-1·50) | 0·41 | | |
| 2-4 | 0·95 (0·55-1·66) | 0·87 | | |
| 5+ | 1·22 (0·51-2·89) | 0·67 | | |
| **Travelled abroad in 2020 [b]** | | | | |
| No | 1 | | | |
| Yes | 1·35 (0·82-2·23) | 0·25 | | |

**Abbreviations:** CI, confidence interval; HELIUS, Healthy Life in an Urban Setting; N.A., not applicable; OR, odds ratio

[a] Those with an equivocal test result were excluded from this analysis [b] Measured at COVID-1 visit (2020) [c] Measured at baseline (2011-2015) [d] Quartiles [e] Presumed higher exposure categories had priority, i.e. if someone was working in a school and as a care worker, they were categorized as a health worker. Caretakers were not included as a category because many had other jobs.

In multivariable analysis, the following variables were removed as they were no longer significant in the multivariable model: migration generation, leaving home to walk the dog or go outside with kids, living with a partner, job setting, living with a child up to 3 years old, number of household members

**Table S6. Factors associated with transitioning to positive SARS-CoV-2 antibody status in African Surinamese participants, Amsterdam, the Netherlands, January 1, 2020 – March 31, 2021**

| | Univariable | | Multivariable | |
|---|---|---|---|---|
| **Characteristic** | **HR (95% CI)** | **P-value** | **aHR (95% CI)** | **P-value** |
| **Sex** | | | | |
| Male | 1 | | | |
| Female | 0·98 (0·62-1·55) | 0·94 | | |
| **Per year increase in age in years** | 0·99 (0·97-1·01) | 0·22 | | |
| **Migration generation** | | | | |
| 1st | 1 | | | |
| 2nd | 1·19 (0·63-2·26) | 0·60 | | |
| **Educational level** [c] | | | | |
| No school/elementary school | 1 | | | |
| Lower vocational/ lower secondary school | 0·60 (0·21-1·76) | 0·36 | | |
| Intermediary vocational/ intermediary secondary school | 0·83 (0·29-2·34) | 0·73 | | |
| Higher vocational/university | 0·49 (0·17-1·44) | 0·20 | | |
| **Labor participation** [c] | | | | |
| Employed | 1 | | | |
| Not in workforce | 1·15 (0·54-2·41) | 0·73 | | |
| Unemployed/on benefits | 1·33 (0·69-2·54) | 0·40 | | |
| Disabled | 0·64 (0·20-2·06) | 0·47 | | |
| Unknown/missing | 1·54 (0·21-11·17) | 0·68 | | |
| **Elementary occupation** [c] | | | | |
| No | 1 | | | |
| Yes | 1·18 (0·81-1·72) | 0·39 | | |
| **Difficulties with Dutch language** | | | | |
| No | 1 | | | |
| Yes | 1·71 (0·94-3·11) | 0·080 | | |
| **Health literacy (SBSQ)** [c] | | | | |
| Adequate | 1 | | | |
| Low | 2·52 (0·79-8·03) | 0·12 | 3·64 (1·08-12·32) | 0·038 |
| **Job setting**[b,e] | | | | |
| No job / caretaker only | 1 | | | |
| Job with no contact within 1·5 meter | 0·46 (0·14-1·57) | 0·22 | | |
| Other job with contact within 1·5 meter | 1·44 (0·80-2·58) | 0·22 | | |
| Child care/schools/higher education | 1·28 (0·58-2·83) | 0·55 | | |
| Bar/restaurant | 0 (0-inf) | 0·88 | | |
| Hospital/long-term care facility/Care worker elsewhere | 1·89 (0·97-3·68) | 0·061 | | |
| **Caretaker** [b] | | | | |
| No | 1 | | | |
| Yes | 1·17 (0·65-2·09) | 0·61 | | |
| **Per 1 person increase in number of household members**[c] | 1·32 (1·16-1·49) | <0·001 | | |
| **Lives with other people** [b] | | | | |
| **Partner** | 1·63 (1·04-2·57) | 0·033 | | |
| **Children up to 3 years old** | 1·67 (0·67-4·13) | 0·27 | | |
| **Children 4 through 12 years old** | 2·04 (1·14-3·65) | 0·017 | | |
| **Children 13 through 17 years old** | 1·77 (0·95-3·29) | 0·070 | | |
| **Children 18+ years old** | 2·16 (1·36-3·43) | 0·001 | 2·31 (1·43-3·71) | <0·001 |
| **Parents or parents-in-law** | 1·03 (0·25-4·21) | 0·97 | | |
| **Other adults** | 2·18 (1·08-4·38) | 0·029 | | |
| **Household member/steady partner with suspected infection**[b] | | | | |

| | | | | |
|---|---|---|---|---|
| N·A·/No | 1 | | 1 | |
| Yes | 4·52 (2·62-7·78) | <0·001 | 4·32 (2·46-7·58) | <0·001 |
| **Number of times left home in the past week [b,d] (per 1 increase)** | 1·01 (0·98-1·04) | 0·4 | | |
| 0-7 | 1 | | | |
| 8-11 | 1·01 (0·55-1·88) | 0·97 | | |
| 12-16 | 0·94 (0·48-1·85) | 0·88 | | |
| 17+ | 1·42 (0·79-2·55) | 0·25 | | |
| **In the past week, left home to [b]:** | | | | |
| **Work** | 1·58 (1·00-2·49) | 0·05 | | |
| **Do groceries** | 0·62 (0·31-1·24) | 0·18 | | |
| **Visit family or friends** | 1·26 (0·80-1·98) | 0·32 | | |
| **Walk the dog or go outside with kids** | 1·42 (0·79-2·54) | 0·24 | | |
| **Walk or exercise outside** | 0·61 (0·39-0·96) | 0·03 | 0·56 (0·35-0·89) | 0·014 |
| **Take care of someone** | 1·06 (0·55-2·07) | 0·87 | | |
| **Pick up prescription medicines or visit doctor** | 1·39 (0·85-2·28) | 0·19 | | |
| **Attend religious service** | 1·81 (0·66-4·96) | 0·25 | | |
| **Visit cultural place** | 0·96 (0·30-3·06) | 0·95 | | |
| **Visit bar or restaurant** | 1·09 (0·64-1·85) | 0·76 | | |
| **Indoor sports** | 1·07 (0·55-2·08) | 0·86 | | |
| **Visit recreational park** | 1·01 (0·56-1·84) | 0·97 | | |
| **Frequency of using public transportation in the past week [b]** | | | | |
| 0 days | 1 | | | |
| 1-2 days | 0·87 (0·50-1·52) | 0·64 | | |
| 3-4 days | 1·06 (0·51-2·18) | 0·89 | | |
| 5-7 days | 1·42 (0·66-3·04) | 0·37 | | |
| **Number of unique visitors at home in the past week [b] (per 1 increase)** | 1·05 (1·00-1·10) | 0·063 | | |
| 0 | 1 | | 1 | |
| 1 | 1·55 (0·84-2·86) | 0·16 | 1·27 (0·66-2·43) | 0·48 |
| 2-4 | 1·79 (1·02-3·14) | 0·041 | 1·80 (1·02-3·20) | 0·044 |
| 5+ | 2·16 (1·01-4·62) | 0·046 | 2·43 (1·12-5·26) | 0·024 |
| **Travelled abroad in 2020 [b]** | | | | |
| No | 1 | | | |
| Yes | 1·20 (0·75-1·93) | 0·45 | | |

**Abbreviations:** CI, confidence interval; HELIUS, Healthy Life in an Urban Setting; N.A., not applicable; OR, odds ratio

[a] Those with an equivocal test result were excluded from this analysis [b] Measured at COVID-1 visit (2020) [c] Measured at baseline (2011-2015) [d] Quartiles [e] Presumed higher exposure categories had priority, i.e. if someone was working in a school and as a care worker, they were categorized as a health worker. Caretakers were not included as a category because many had other jobs.

In multivariable analysis, the following variables were removed as they were no longer significant in the multivariable model: Dutch language skills, leaving home to work in the past week, number of visitors (continuous variable), living with a child of 13 through 17 years old, leaving home to do groceries in the past week, living with a partner, number of household members, leaving with other adults, living with a child of 4 through 12 years old,  job setting.

**Table S7. Factors associated with transitioning to positive SARS-CoV-2 antibody status in Ghanaian participants, Amsterdam, the Netherlands, January 1, 2020 – March 31, 2021**

| | Univariable | | Multivariable | |
|---|---|---|---|---|
| **Characteristic** | **HR (95% CI)** | **P-value** | **aHR (95% CI)** | **P-value** |
| **Sex** | | | | |
| Male | 1 | | | |
| Female | 1·14 (0·82-1·59) | 0·46 | | |
| **Per year increase in age in years** | 1·01 (0·99-1·03) | 0·39 | | |
| **Migration generation** | | | | |
| 1st | 1 | | | |
| 2nd | 0·38 (0·05-2·73) | 0·34 | | |
| **Educational level** [c] | | | | |
| No school/elementary school | 1 | | | |
| Lower vocational/ lower secondary school | 0·88 (0·58-1·33) | 0·55 | | |
| Intermediary vocational/ intermediary secondary school | 0·83 (0·51-1·34) | 0·45 | | |
| Higher vocational/university | 0·72 (0·36-1·46) | 0·37 | | |
| **Labor participation** [c] | | | | |
| Employed | 1 | | | |
| Not in workforce | 0·64 (0·20-2·02) | 0·45 | | |
| Unemployed/on benefits | 1·10 (0·72-1·69) | 0·66 | | |
| Disabled | 0·92 (0·49-1·73) | 0·81 | | |
| Unknown/missing | 0·54 (0·13-2·22) | 0·40 | | |
| **Elementary occupation** [c] | | | | |
| No | 1 | | | |
| Yes | 0·97 (0·76-1·23) | 0·80 | | |
| **Difficulties with Dutch language** | | | | |
| No | 1 | | | |
| Yes | 1·17 (0·69-1·97) | 0·57 | | |
| **Health literacy (SBSQ)** [c] | | | | |
| Adequate | 1 | | | |
| Low | 1·06 (0·81-1·37) | 0·69 | | |
| **Job setting**[b,e] | | | | |
| No job / caretaker only | 1 | | | |
| Job with no contact within 1·5 meter | 1·10 (0·67-1·82) | 0·72 | | |
| Other job with contact within 1·5 meter | 1·21 (0·79-1·87) | 0·39 | | |
| Child care/schools/higher education | 1·88 (0·78-4·53) | 0·16 | | |
| Bar/restaurant | 1·73 (0·91-3·29) | 0·097 | | |
| Hospital/long-term care facility/Care worker elsewhere | 1·02 (0·50-2·07) | 0·96 | | |
| **Caretaker** [b] | | | | |
| No | 1 | | | |
| Yes | 0·80 (0·37-1·71) | 0·57 | | |
| **Per 1 person increase in number of household members**[c] | 1·09 (0·98-1·22) | 0·12 | | |
| **Lives with other people** [b] | | | | |
| **Partner** | 0·86 (0·61-1·21) | 0·40 | | |
| **Children up to 3 years old** | 1·46 (0·84-2·55) | 0·18 | | |
| **Children 4 through 12 years old** | 0·99 (0·67-1·45) | 0·96 | | |
| **Children 13 through 17 years old** | 1·25 (0·86-1·81) | 0·24 | | |
| **Children 18+ years old** | 1·25 (0·89-1·75) | 0·20 | | |
| **Parents or parents-in-law** | 1·18 (0·44-3·22) | 0·75 | | |
| **Other adults** | 1·31 (0·87-1·97) | 0·19 | | |
| **Household member/steady partner with suspected infection**[b] | | | | |

| | | | | |
|---|---|---|---|---|
| N·A·/No | 1 | | | |
| Yes | 0·72 (0·29-1·76) | 0·48 | | |
| **Number of times left home in the past week [b,d] (per 1 increase)** | 1·00 (0·97-1·04) | 0·85 | | |
| 0-7 | 1 | | | |
| 8-11 | 0·84 (0·57-1·24) | 0·39 | | |
| 12-16 | 0·87 (0·54-1·40) | 0·57 | | |
| 17+ | 1·19 (0·67-2·13) | 0·56 | | |
| **In the past week, left home to [b]:** | | | | |
| **Work** | 1·41 (1·00-2·00) | 0·050 | 1·49 (1·05-2·11) | 0·024 |
| **Do groceries** | 1·01 (0·58-1·76) | 0·98 | | |
| **Visit family or friends** | 0·93 (0·63-1·37) | 0·74 | | |
| **Walk the dog or go outside with kids** | 1·66 (0·93-2·96) | 0·084 | 1·90 (1·06-3·41) | 0·031 |
| **Walk or exercise outside** | 0·86 (0·61-1·20) | 0·38 | | |
| **Take care of someone** | 1·44 (0·70-2·95) | 0·33 | | |
| **Pick up prescription medicines or visit doctor** | 0·96 (0·64-1·45) | 0·86 | | |
| **Attend religious service** | 1·50 (1·08-2·09) | 0·017 | | |
| **Visit cultural place** | 1·08 (0·15-7·79) | 0·94 | | |
| **Visit bar or restaurant** | 0·42 (0·15-1·13) | 0·086 | 0·34 (0·12-0·92) | 0·034 |
| **Indoor sports** | 1·31 (0·75-2·29) | 0·35 | | |
| **Visit recreational park** | 1·03 (0·51-2·12) | 0·93 | | |
| **Frequency of using public transportation in the past week [b]** | | | | |
| 0 days | 1 | | | |
| 1-2 days | 0·95 (0·60-1·51) | 0·84 | | |
| 3-4 days | 1·24 (0·73-2·09) | 0·43 | | |
| 5-7 days | 1·06 (0·71-1·60) | 0·78 | | |
| **Number of unique visitors at home in the past week [b] (per 1 increase)** | 1·05 (0·96-1·15) | 0·39 | | |
| 0 | 1 | | | |
| 1 | 0·78 (0·46-1·33) | 0·38 | | |
| 2-4 | 1·29 (0·81-2·05) | 0·29 | | |
| 5+ | 0·69 (0·17-2·80) | 0·61 | | |
| **Travelled abroad in 2020 [b]** | | | | |
| No | 1 | | | |
| Yes | 0·85 (0·56-1·28) | 0·44 | | |

**Abbreviations:** CI, confidence interval; HELIUS, Healthy Life in an Urban Setting; N.A., not applicable; OR, odds ratio

[a] Those with an equivocal test result were excluded from this analysis [b] Measured at COVID-1 visit (2020) [c] Measured at baseline (2011-2015) [d] Quartiles [e] Presumed higher exposure categories had priority, i.e. if someone was working in a school and as a care worker, they were categorized as a health worker. Caretakers were not included as a category because many had other jobs.

In multivariable analysis, the following variables were removed as they were no longer significant in the multivariable model: number of household members, job setting, living with a child up to 3 years old, living with an adult child, living with other adults, attending a religious service.

**Table S8. Factors associated with transitioning to positive SARS-CoV-2 antibody status in Turkish participants, Amsterdam, the Netherlands, January 1, 2020 – March 31, 2021**

| | Univariable | | Multivariable | |
|---|---|---|---|---|
| **Characteristic** | **HR (95% CI)** | **P-value** | **aHR (95% CI)** | **P-value** |
| **Sex** | | | | |
| Male | 1 | | | |
| Female | 1·01 (0·68-1·49) | 0·98 | | |
| **Per year increase in age in years** | 0·99 (0·98-1·01) | 0·45 | | |
| **Migration generation** | | | | |
| 1st | 1 | | | |
| 2nd | 1·13 (0·74-1·74) | 0·58 | | |
| **Educational level** [c] | | | | |
| No school/elementary school | 1 | | 1 | |
| Lower vocational/ lower secondary school | 0·61 (0·33-1·10) | 0·098 | 0·56 (0·31-1·02) | 0·059 |
| Intermediary vocational/ intermediary secondary school | 0·58 (0·34-0·98) | 0·043 | 0·57 (0·33-0·99) | 0·044 |
| Higher vocational/university | 0·54 (0·31-0·94) | 0·028 | 0·48 (0·27-0·85) | 0·011 |
| **Labor participation** [c] | | | | |
| Employed | 1 | | | |
| Not in workforce | 1·05 (0·60-1·85) | 0·87 | | |
| Unemployed/on benefits | 0·85 (0·48-1·51) | 0·59 | | |
| Disabled | 1·13 (0·51-2·47) | 0·78 | | |
| Unknown/missing | 0·11 (0·00-84·89) | 0·53 | | |
| **Elementary occupation** [c] | | | | |
| No | 1 | | | |
| Yes | 1·14 (0·90-1·44) | 0·28 | | |
| **Difficulties with Dutch language** | | | | |
| No | 1 | | | |
| Yes | 1·17 (0·78-1·74) | 0·46 | | |
| **Health literacy (SBSQ)** [c] | | | | |
| Adequate | 1 | | | |
| Low | 1·24 (0·87-1·77) | 0·23 | | |
| **Job setting**[b,e] | | | | |
| No job / caretaker only | 1 | | | |
| Job with no contact within 1·5 meter | 1·03 (0·57-1·85) | 0·93 | | |
| Other job with contact within 1·5 meter | 1·00 (0·62-1·61) | 1·00 | | |
| Child care/schools/higher education | 0·73 (0·29-1·87) | 0·52 | | |
| Bar/restaurant | 0·64 (0·09-4·61) | 0·67 | | |
| Hospital/long-term care facility/Care worker elsewhere | 1·22 (0·63-2·37) | 0·57 | | |
| **Caretaker** [b] | | | | |
| No | 1 | | 1 | |
| Yes | 1·63 (0·93-2·88) | 0·089 | 1·93 (1·08-3·45) | 0·027 |
| **Per 1 person increase in number of household members**[c] | 1·23 (1·06-1·42) | 0·006 | 1·17 (1·01-1·36) | 0·041 |
| **Lives with other people** [b] | | | | |
| **Partner** | 1·50 (0·97-2·32) | 0·067 | | |
| **Children up to 3 years old** | 1·03 (0·55-1·93) | 0·93 | | |
| **Children 4 through 12 years old** | 0·98 (0·60-1·58) | 0·93 | | |
| **Children 13 through 17 years old** | 1·42 (0·90-2·25) | 0·13 | | |
| **Children 18+ years old** | 1·43 (0·97-2·11) | 0·073 | | |
| **Parents or parents-in-law** | 0·87 (0·40-1·87) | 0·73 | | |
| **Other adults** | 0·77 (0·28-2·10) | 0·63 | | |

| | | | | |
|---|---|---|---|---|
| **Household member/steady partner with suspected infection[b]** | | | | |
| N·A·/No | 1 | | 1 | |
| Yes | 2·17 (1·30-3·62) | 0·003 | 2·19 (1·29-3·74) | 0·004 |
| **Number of times left home in the past week [b,d] (per 1 increase)** | 1·00 (0·97-1·03) | 0·98 | | |
| 0-7 | 1 | | | |
| 8-11 | 0·92 (0·54-1·59) | 0·79 | | |
| 12-16 | 0·97 (0·57-1·65) | 0·91 | | |
| 17+ | 0·88 (0·51-1·53) | 0·67 | | |
| **In the past week, left home to [b]:** | | | | |
| **Work** | 1·31 (0·89-1·94) | 0·17 | | |
| **Do groceries** | 1·11 (0·59-2·08) | 0·75 | | |
| **Visit family or friends** | 1·16 (0·78-1·72) | 0·46 | | |
| **Walk the dog or go outside with kids** | 1·25 (0·78-2·00) | 0·40 | | |
| **Walk or exercise outside** | 1·02 (0·68-1·54) | 0·92 | | |
| **Take care of someone** | 1·42 (0·76-2·66) | 0·28 | | |
| **Pick up prescription medicines or visit doctor** | 1·13 (0·70-1·81) | 0·63 | | |
| **Attend religious service** | 1·46 (0·89-2·39) | 0·13 | | |
| **Visit cultural place** | 1·08 (0·40-2·93) | 0·90 | | |
| **Visit bar or restaurant** | 0·85 (0·56-1·32) | 0·49 | | |
| **Indoor sports** | 1·29 (0·72-2·31) | 0·40 | | |
| **Visit recreational park** | 1·13 (0·69-1·84) | 0·64 | | |
| **Frequency of using public transportation in the past week [b]** | | | | |
| 0 days | 1 | | | |
| 1-2 days | 1·07 (0·65-1·76) | 0·80 | | |
| 3-4 days | 1·43 (0·52-3·91) | 0·50 | | |
| 5-7 days | 0·88 (0·28-2·79) | 0·84 | | |
| **Number of unique visitors at home in the past week [b] (per 1 increase)** | 0·98 (0·90-1·06) | 0·55 | | |
| 0 | 1 | | | |
| 1 | 1·05 (0·56-1·98) | 0·89 | | |
| 2-4 | 1·25 (0·80-1·94) | 0·34 | | |
| 5+ | 0·77 (0·35-1·70) | 0·53 | | |
| **Travelled abroad in 2020 [b]** | | | | |
| No | 1 | | | |
| Yes | 1·16 (0·79-1·71) | 0·46 | | |

**Abbreviations:** CI, confidence interval; HELIUS, Healthy Life in an Urban Setting; N.A., not applicable; OR, odds ratio

[a] Those with an equivocal test result were excluded from this analysis [b] Measured at COVID-1 visit (2020) [c] Measured at baseline (2011-2015) [d] Quartiles [e] Presumed higher exposure categories had priority, i.e. if someone was working in a school and as a care worker, they were categorized as a health worker. Caretakers were not included as a category because many had other jobs.

In multivariable analysis, the following variables were removed as they were no longer significant in the multivariable model: living with a partner, living with an adult child, living with a child aged 13 through 17 years old, attending a religious service, number of unique visitors at home in the past week, living with a child up to 3 years old, leaving home to work in the past week.

**Table S9. Factors associated with transitioning to positive SARS-CoV-2 antibody status in Moroccan participants, Amsterdam, the Netherlands, January 1, 2020 – March 31, 2021**

| | Univariable | | Multivariable | |
|---|---|---|---|---|
| **Characteristic** | **HR (95% CI)** | **P-value** | **aHR (95% CI)** | **P-value** |
| **Sex** | | | | |
| Male | 1 | | | |
| Female | 1·16 (0·79-1·70) | 0·47 | | |
| **Per year increase in age in years** | 1·00 (0·98-1·01) | 0·63 | | |
| **Migration generation** | | | | |
| $1^{st}$ | 1 | | | |
| $2^{nd}$ | 1·05 (0·68-1·62) | 0·84 | | |
| **Educational level** [c] | | | | |
| No school/elementary school | 1 | | | |
| Lower vocational/ lower secondary school | 1·24 (0·70-2·20) | 0·48 | | |
| Intermediary vocational/ intermediary secondary school | 0·96 (0·57-1·62) | 0·89 | | |
| Higher vocational/university | 0·71 (0·40-1·27) | 0·25 | | |
| **Labor participation** [c] | | | | |
| Employed | 1 | | | |
| Not in workforce | 1·21 (0·72-2·05) | 0·48 | | |
| Unemployed/on benefits | 0·91 (0·50-1·65) | 0·76 | | |
| Disabled | 1·01 (0·44-2·34) | 0·98 | | |
| Unknown/missing | 3·60 (1·09-11·92) | 0·035 | | |
| **Elementary occupation** [c] | | | | |
| No | 1 | | | |
| Yes | 1·09 (0·87-1·36) | 0·48 | | |
| **Difficulties with Dutch language** | | | | |
| No | 1 | | | |
| Yes | 1·12 (0·76-1·67) | 0·57 | | |
| **Health literacy (SBSQ)** [c] | | | | |
| Adequate | 1 | | | |
| Low | 1·18 (0·86-1·62) | 0·32 | | |
| **Job setting**[b,e] | | | | |
| No job / caretaker only | 1 | | | |
| Job with no contact within 1·5 meter | 1·11 (0·63-1·95) | 0·74 | | |
| Other job with contact within 1·5 meter | 0·65 (0·39-1·09) | 0·10 | | |
| Child care/schools/higher education | 1·20 (0·67-2·13) | 0·55 | | |
| Bar/restaurant | 0·93 (0·22-3·86) | 0·92 | | |
| Hospital/long-term care facility/Care worker elsewhere | 0·95 (0·47-1·91) | 0·89 | | |
| **Caretaker** [b] | | | | |
| No | 1 | | | |
| Yes | 1·15 (0·68-1·93) | 0·62 | | |
| **Per 1 person increase in number of household members**[c] | 1·20 (1·08-1·34) | 0·001 | 1·22 (1·09-1·36) | <0·001 |
| **Lives with other people** [b] | | | | |
| **Partner** | 1·10 (0·73-1·66) | 0·65 | | |
| **Children up to 3 years old** | 1·30 (0·75-2·25) | 0·36 | | |
| **Children 4 through 12 years old** | 0·70 (0·44-1·12) | 0·13 | 0·53 (0·33-0·87) | 0·012 |
| **Children 13 through 17 years old** | 1·72 (1·17-2·54) | 0·006 | | |
| **Children 18+ years old** | 2·01 (1·37-2·95) | <0·000 | | |
| **Parents or parents-in-law** | 0·90 (0·40-2·06) | 0·82 | | |
| **Other adults** | 1·01 (0·44-2·31) | 0·98 | | |

| | | | | |
|---|---|---|---|---|
| **Household member/steady partner with suspected infection[b]** | | | | |
| N·A·/No | 1 | | 1 | |
| Yes | 2·45 (1·56-3·85) | <0·000 | 2·54 (1·58-4·08) | <0·001 |
| **Number of times left home in the past week [b,d] (per 1 increase)** | 0·99 (0·96-1·01) | 0·37 | | |
| 0-7 | 1 | | | |
| 8-11 | 0·75 (0·44-1·30) | 0·31 | | |
| 12-16 | 0·70 (0·41-1·20) | 0·20 | | |
| 17+ | 0·80 (0·49-1·32) | 0·39 | | |
| **In the past week, left home to [b]:** | | | | |
| **Work** | 0·67 (0·45-1·00) | 0·048 | | |
| **Do groceries** | 0·63 (0·33-1·17) | 0·15 | 0·45 (0·24-0·87) | <0·001 |
| **Visit family or friends** | 1·20 (0·81-1·77) | 0·37 | | |
| **Walk the dog or go outside with kids** | 0·84 (0·52-1·37) | 0·50 | | |
| **Walk or exercise outside** | 0·77 (0·52-1·15) | 0·20 | | |
| **Take care of someone** | 0·92 (0·53-1·58) | 0·77 | | |
| **Pick up prescription medicines or visit doctor** | 0·90 (0·57-1·43) | 0·67 | | |
| **Attend religious service** | 1·27 (0·68-2·37) | 0·47 | | |
| **Visit cultural place** | 0·38 (0·09-1·54) | 0·18 | | |
| **Visit bar or restaurant** | 1·03 (0·69-1·53) | 0·91 | | |
| **Indoor sports** | 0·94 (0·50-1·75) | 0·85 | | |
| **Visit recreational park** | 1·00 (0·64-1·56) | 1·00 | | |
| **Frequency of using public transportation in the past week [b]** | | | | |
| 0 days | 1 | | | |
| 1-2 days | 0·56 (0·33-0·95) | 0·031 | | |
| 3-4 days | 1·32 (0·66-2·64) | 0·44 | | |
| 5-7 days | 0·43 (0·14-1·38) | 0·16 | | |
| **Number of unique visitors at home in the past week [b] (per 1 increase)** | 1·04 (0·99-1·11) | 0·14 | | |
| 0 | 1 | | 1 | |
| 1 | 0·42 (0·18-0·97) | 0·042 | 0·37 (0·16-0·88) | 0·023 |
| 2-4 | 1·02 (0·66-1·59) | 0·92 | 0·85 (0·53-1·36) | 0·51 |
| 5+ | 1·42 (0·77-2·59) | 0·26 | 1·28 (0·67-2·43) | 0·47 |
| **Travelled abroad in 2020 [b]** | | | | |
| No | 1 | | | |
| Yes | 0·96 (0·65-1·41) | 0·83 | | |

**Abbreviations:** CI, confidence interval; HELIUS, Healthy Life in an Urban Setting; N.A., not applicable; OR, odds ratio

[a] Those with an equivocal test result were excluded from this analysis [b] Measured at COVID-1 visit (2020) [c] Measured at baseline (2011-2015) [d] Quartiles [e] Presumed higher exposure categories had priority, i.e. if someone was working in a school and as a care worker, they were categorized as a health worker. Caretakers were not included as a category because many had other jobs.

In multivariable analysis, the following variables were removed as they were no longer significant in the multivariable model: number of visitors (continuous), attending a cultural place in the past week, living with an adult child, labor participation, living with a child aged 13 through 17 years, job setting, leaving home to work in the past week, frequency of using public transportation in the past week.
